# Leveraging genome-wide effects on gene expression to identify disease-critical genes with *trans*-genetic components

**DOI:** 10.64898/2026.02.23.26346922

**Authors:** Krishna Brunton, Michelle Franc Ragsac, Tiffany Amariuta

## Abstract

Genome-wide association studies (GWAS) have implicated tens of thousands of genetic variants associated with complex traits and polygenic diseases. Colocalizing GWAS variants with variants that may regulate gene expression, via expression quantitative trait loci (eQTL) mapping, has successfully led to the identification of disease-critical genes and their cell types of action. Recent studies predominantly colocalize proximal *cis*-eQTLs, which are estimated to regulate ∼10% of variance in gene expression levels. However, *trans*-eQTLs have been hypothesized to account for an additional ∼20% of expression levels, although few studies have attempted to quantify the variance explained by empirically associated *trans*-eQTLs. Here, we introduce EGRET (Estimating Genome-wide Regulatory Effects on the Transcriptome), an ensemble framework that jointly models *cis*-eQTLs with three distinct *trans*-eQTL mapping approaches: standard pairwise association testing via Matrix eQTL, and two functionally-informed methods, trans-PCO and GBAT. In real data, EGRET produced 353,408 predictive gene expression models (cross-validation R^2^ > 0, p < 0.01) across 49 GTEx tissues, including 12,317 gene-tissue pairs with a significantly nonzero *trans*-heritable component. For this set of genes, EGRET models explain 33% more gene expression variance than *cis*-eQTL models (EGRET average R^2^ = 0.104, FUSION average R^2^ = 0.078). We found that putative trans-regulating variants of EGRET models are enriched for regulatory elements such as enhancers, histone marks, and *cis*-eQTLs of other genes. We then hypothesized that EGRET models could nominate new disease-critical genes via a transcriptome-wide association study (TWAS) framework that models genome-wide regulatory effects on gene expression. In simulations of theoretically representative gene expression architectures (∼30% heritability, where more than 70% is distal), EGRET increased the power to detect disease-critical genes by 1.2x-3.1x compared to *cis*-eQTL models. In real data analysis, we identified disease-associated genes via TWAS across GWAS summary statistics for 78 complex traits and polygenic diseases using gene expression prediction models from EGRET, *cis*-eQTL FUSION, and two state-of-the-art *trans*-eQTL TWAS methods, MOSTWAS and BGW-TWAS. EGRET identified 450,825 gene-disease associations that were not identified by FUSION models, 2,900 associations not identified by MOSTWAS, and 5,498 associations not identified by BGW-TWAS. Finally, we used EGRET models to construct gene regulatory networks, some of which harbored genes that were jointly associated with complex traits. For example, the gene members of the network defined by *ARHGEF3*, whose *cis*-regulatory variants help predict expression of 10 genes in *trans*, were concordantly associated with platelet count using EGRET but not FUSION models. Overall, we find that modeling the genome-wide genetic component of gene expression greatly boosts the detection of disease-critical genes and helps define gene regulatory networks while improving the characterization of GWAS variants.

## Introduction

Genome-wide association studies (GWAS) identify associations between disease status and sites of genetic variation across individuals. Most of these associations implicate noncoding regulatory variants, which are largely thought to act through gene expression regulation^1–6^. *Cis*-eQTLs (expression quantitative trait loci) are variants that tend to have the strongest nearby effects on gene expression, i.e., within 500 kilobase pairs (kb)^7,8^. However, an average of only 30% of the genetic variation impacting a gene’s expression level, referred to as heritability (*h^2^*), can be explained by *cis*-eQTLs^9,10^. On the other hand, the remaining heritability is likely to be explained by *trans*-eQTLs, variants more than 500 kb from the target gene or even residing on different chromosomes^11–13^. Notably, *trans*-regulatory effects have been shown to be disproportionately enriched for disease-associated loci^12,14,15^. For example, GWAS loci for systemic lupus erythematosus (SLE) colocalize with *trans*-eQTLs of *IKZF1* target genes^8,14^; *IKZF1* is a transcription factor that drives the differentiation of B and T cells. Although not quantified in human studies, in yeast, upwards of 77% of heritability across 46 growth traits is captured within *trans*-eQTL hotspots^16^.

While the power of *trans*-eQTL discovery is limited by the sample size of eQTL cohorts, several statistical approaches have been previously developed to reduce the dimensionality and testing burden. Approaches such as ARCHIE, Teejas, and *trans*-PCO test individual variants against sets of genes^15,17,18^, while the Gene-Based Association Test (GBAT) method identifies *trans*-acting regulatory genes^19^. While these approaches can more powerfully identify variants that are significantly associated with expression changes across the transcriptome, there has been little progress in evaluating how much of the missing heritability of gene expression can be explained by these *trans*-eQTLs. A framework that evaluates the proportion of gene expression variance explained by *trans*-eQTLs would enable direct quantitative comparison of different eQTL mapping methods and could improve gene expression imputation in GWAS cohorts for transcriptome-wide association studies (TWAS), which historically colocalize *cis*-eQTLs with disease loci to identify disease-critical genes^4,20^. Two approaches, BGW-TWAS and MOSTWAS, currently aim to address this gap^21,22^, but include a limited set of feature selection methods that may be susceptible to high false positive rates (FPRs) due to unaccounted cross-mappable regions^23^.

In this study, we developed EGRET (**E**stimating **G**enome-wide **R**egulatory **E**ffects on the **T**ranscriptome), a multivariate linear model designed to identify genomic loci that are predictive of gene expression levels. EGRET integrates predictions from existing *trans*-regulatory loci mapping approaches (Matrix eQTL^24^, GBAT^19^, and *trans*-PCO^17^) and determines the optimal weighted combination of regulatory variants that best explains gene expression. We also develop EGRET-TWAS, a genome-wide summary statistics-based TWAS framework that identifies novel gene-disease associations.

We performed extensive simulations to model the genetic regulation of gene expression and assess the value of EGRET in improving expression prediction models and power to identify disease-critical genes in TWAS. We then used 49 tissue-specific genotype/gene expression datasets from GTEx^25^ to benchmark EGRET prediction models against *cis*-eQTL models generated with FUSION^4^ and constituent *trans*-eQTL models generated separately by Matrix eQTL, GBAT, and *trans*-PCO. We additionally evaluated the biological plausibility of *trans*-eQTL loci detected by EGRET models via functional enrichment of model weights and network analysis of tissue-specific and nonspecific *trans*-regulating genes using established mechanisms from previous studies. Lastly, we benchmarked EGRET gene models in a TWAS framework against MOSTWAS and BGW-TWAS^21,22^, two recently published methods that integrate distal regulatory variants into gene expression models and perform colocalization with GWAS summary statistics. For this analysis, we investigated 78 independent complex traits and polygenic diseases and only considered genes that are not cross-mappable, as these sequences have been previously shown to produce false positives in *trans*-eQTL mapping^23^ and therefore might bias TWAS results if including *trans*-eQTLs in gene expression prediction models.

Overall, we find that modeling genome-wide regulatory effects on gene expression (1) helps account for a significant proportion of gene expression regulation not explained by *cis*-eQTLs although much less than optimistically suggested by previous studies, (2) implicates new genes in the biology of human disease and complex traits, (3) greatly improves the mechanistic interpretation of GWAS variants, and (4) helps construct tissue-specific gene regulatory networks based on co-regulation patterns. We attribute these new findings to our multi-stage feature selection strategy and reduction of false positives by omitting highly cross-mappable regions with other genes.

## Results

### Overview of EGRET

EGRET aims to boost the power of TWAS to nominate relevant disease associated genes by incorporating *trans*-eQTLs to improve the accuracy of gene expression models (**Figure 1A**). *Trans*-eQTLs may act on their target gene through several mechanisms, such as *cis*-regulation of a transcription factor gene that subsequently modulates downstream targets in a cell-type-specific manner^26^, regulation of a lincRNA capable of acting on multiple genes in *cis* or *trans*^27,28^, or perturbation of a gene encoding a signaling molecule, such as a receptor, thereby influencing cell-cell communication and coordinated pathway-level gene expression^29,30^.

**Figure 1.**
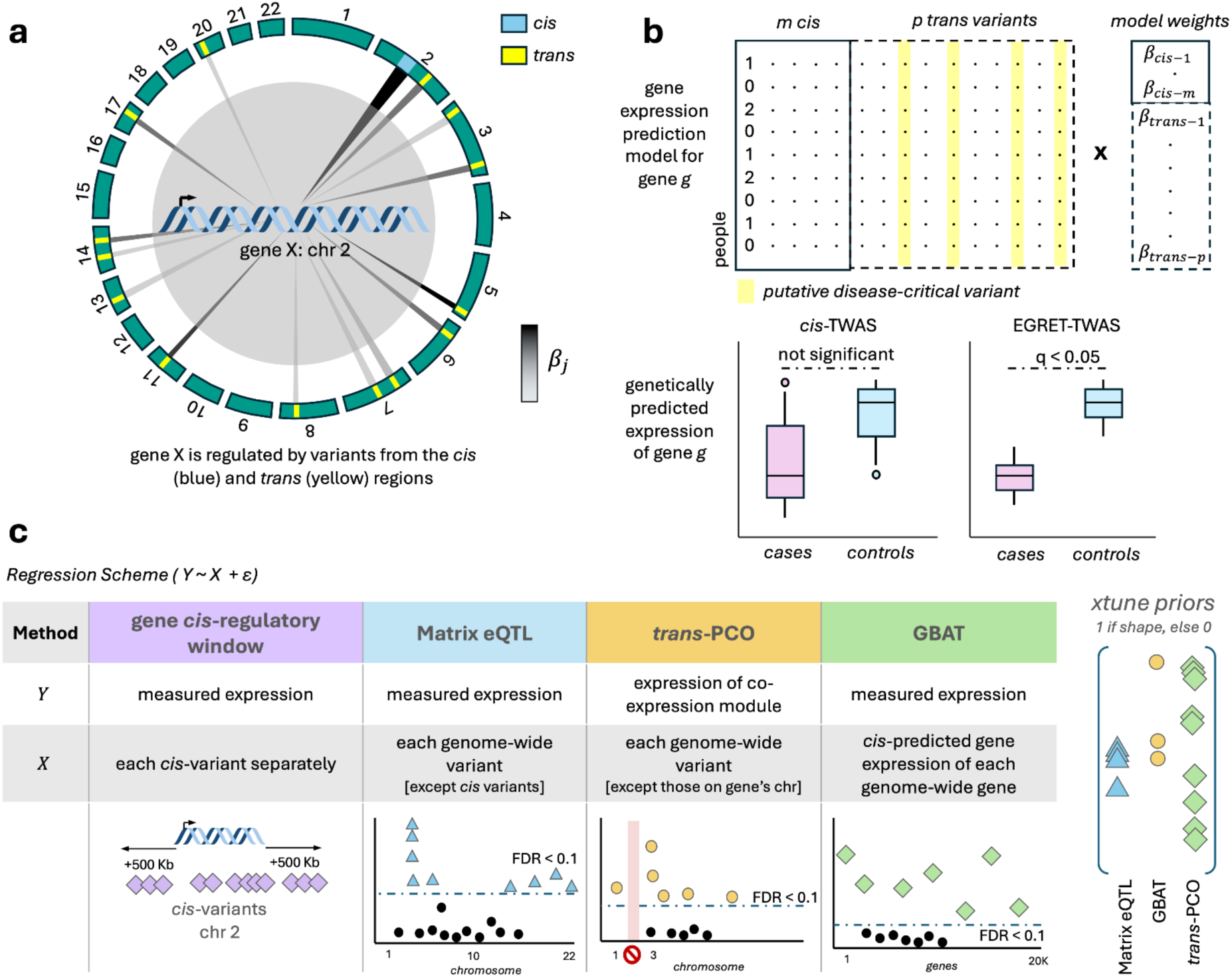
EGRET overview. (a) Effects on gene expression originate from local (*cis*) and distal (*trans*) regions. Incorporating genome-wide eQTLs into gene expression models may enhance our understanding of transcriptional regulation. *β_j_* symbolically represents the relative effect sizes of *cis*- and *trans*-eQTLs. (b) Joint modeling of both *cis* and *trans* variants within a multivariate regression framework has the potential to explain more gene expression variance compared to models that consider only *cis* variants. Accounting for *trans*-regulatory effects may also enhance the ability to identify disease-critical genes influenced by distal genetic variation. (c) (Left) EGRET employs three complementary high-dimensional feature selection strategies to identify candidate regulatory variants across the genome in addition to modeling *cis*-variants. (Right) The xtune LASSO sparse multivariate regression framework integrates prior information as categorical uniform priors to improve prediction performance. Abbreviations: chr, chromosome.

Because traditional eQTL mapping approaches such as pairwise testing have limited power to detect *trans*-eQTLs due to small effect sizes, large testing burdens, and poor reproducibility, EGRET integrates three orthogonal *trans*-regulatory loci mapping methods, Matrix eQTL, GBAT, and *trans*-PCO, to identify putative *trans*-acting elements and optimally incorporate them into gene expression prediction models (**Figure 1B**).

The choice of these three methods reflects three orthogonal approaches to *trans*-regulatory loci mapping. The most direct approach for *trans*-eQTL mapping is to conduct association tests across every variant-gene pair which often is very computationally intensive. Therefore, we first use Matrix eQTL, an efficient method to identify *trans*-eQTLs with large effect sizes due to its univariate regression framework^24^. Second, *cis-*genetic regulation of some genes may cause changes in expression of downstream target genes. GBAT captures these effects by nominating upstream genes whose predicted *cis*-expression shows a significant association with the measured expression of the target gene, indicating a substantial regulatory effect^19^. Third, to find highly prevalent *trans*-regulators that act on many genes simultaneously, *trans*-PCO implements six principal component (PC)-based multivariate association tests to discover putative *trans*-eQTLs that regulate modules of co-expressed genes^17^. Because of the conceptual similarity of their approaches, we elected to integrate only *trans*-PCO, as opposed to ARCHIE or Teejas into the EGRET framework. EGRET implements these tools in an integrated framework for gene expression prediction (**Methods**). We also model standard confounders, such as genotype PCs to capture population structure, expression PCs to capture global expression changes across many genes, as well as recorded batch effects such as age and sex. Finally, we determine significant variant-gene associations based on a 10% false discovery rate (FDR) applied across the number of elements tested: the number of single nucleotide polymorphisms (SNPs) for Matrix eQTL and *trans*-PCO or the number of genes for GBAT. The LASSO weighted variants of the upstream *cis*-models used for GBAT are used as putative *trans*-eQTLs of the target gene. Modeling these variants jointly with *cis*-variants within 1 megabase (1 Mb) of the gene’s transcription start site (TSS) allows EGRET to prioritize variants genome-wide that may be critical for improving gene expression models (**Figure 1C**).

To best capture nearby and distal effects that may operate through different genetic architectures, EGRET then fits four multivariate regression methods to *cis*-variants and associated *trans*-variants from Matrix eQTL, GBAT, and *trans*-PCO. Previous work recognized that sparse regression models (e.g., LASSO and elastic net (enet)) may capture distinct genetic architectures of expression regulation compared to linear mixed-models (e.g., Best Linear Unbiased Prediction (BLUP)^4,20^). Therefore, EGRET employs standard LASSO, elastic net, and BLUP to genome-wide SNPs with 5-fold cross validation (CV). Additionally, EGRET leverages xtune LASSO^31^, an empirical Bayes approach to learn group priors of *trans*-eQTL mapping approaches, helping EGRET infer the most important features. In addition to genotype and expression data, our input to xtune LASSO includes a binary matrix of model features (SNPs) by each *trans*-eQTL mapping method; variants identified by each method are given a value of 1 to indicate membership. xtune LASSO computes a posterior probability proportional to the predictive power of features from each mapping method (**Methods**). Moreover, to accommodate the fact that the proportions of *cis*- vs. *trans*-genetic regulation will vary greatly across genes, for each regularization scheme, we additionally calculate the CV R^2^ for different sets of variants (*cis*, *trans*, and genome-wide), discussed in detail below. Across these model variations, EGRET selects the model with the greatest CV R^2^ for downstream analyses, including TWAS and constructing gene regulatory networks.

While highly cross-mappable genomic regions present bias in studies involving read mapping data (i.e., RNA-sequencing), the risk for false positive associations is especially high in *trans*-eQTL studies^23^. This is because when reads are mapped promiscuously to multiple genes with similar sequence content, this can make a *cis*-eQTL of one gene appear erroneously as a *trans*-eQTL of many genes and may introduce false gene regulatory networks. Therefore, in an effort to draw meaningful conclusions from benchmarking EGRET against MOSTWAS and BGW-TWAS in terms of gene model construction and gene-disease association via TWAS, we implemented a stringent cross-mappability pruning procedure. Highly cross-mappable gene pairs were identified based on an empirically defined background rate of misaligned reads mapping to the target gene (**Methods**). For gene pairs exceeding this threshold, we excluded genetic variants located within a 100 kb window centered on the transcription start site (TSS) of the affected gene to reduce the likelihood of spurious *trans*-eQTL signals. Alternative thresholds were evaluated, including 0 misaligned reads, 4 misaligned reads, and gene-specific background estimates (**Supplementary Figure 1**), but distributions of gene model performance across EGRET and FUSION were robust to this choice of threshold.

### Simulations

In order to understand the degree to which *trans*-eQTL effects can be captured by regularized multivariate linear models, we performed extensive simulations of varying realistic gene expression architectures for which we constructed simplified EGRET models, which we refer to as Matrix eQTL-only EGRET below. Due to the complex nature of gene co-expression and inherent difficulty in its realistic simulation, we restricted simulations to Matrix eQTL associations and excluded potential contributions from *trans*-PCO and GBAT methods. As this simplification precludes a fair comparison of EGRET with other *trans*-TWAS methods, we benchmarked EGRET against published MOSTWAS and BGW-TWAS performed exclusively in real data. In regards to this, these simulations are intended to provide a conservative baseline for assessing the benefits of the more comprehensive EGRET model. Specifically, we evaluated how well linear models capture the effects of *cis*- and *trans*-eQTLs when jointly modeled and how genome-wide gene expression models perform at nominating disease-critical genes compared to models that include only *cis*-variants, such as those generated by FUSION^4^.

To evaluate the first point, we randomly sampled European-ancestry individuals from the Genotype-Tissue Expression (GTEx) cohort to leverage individual-level SNP genotyping data^7^. By default, we limited our analyses to 715 European-ancestry individuals to minimize confounding due to population structure. For each individual, we simulated gene expression using variants across the genome that we selected to be causal and preset the total heritability (or variance explained) from *cis*- and *trans*-eQTLs (**Methods**). Causal effect sizes for *cis*- and *trans*-eQTLs were sampled separately from normal distributions with mean 0 and variance equal to the per SNP *cis-* or *trans*-heritability. Per SNP heritability was calculated as the total *cis-* or *trans*-heritability divided by the number of *cis*- or *trans*-eQTLs for a given gene, respectively, where the default is 5 for *cis*-eQTLs and 20 for *trans*-eQTLs. Consequently, effect sizes for *trans*-eQTLs were smaller on average than *cis*-eQTLs, consistent with our current understanding of relative eQTL effect sizes^12^. Non-causal SNPs were assigned effect sizes of 0. Environmental noise was sampled from a normal distribution with mean 0 and variance equal to one minus the empirical heritability of the simulated gene’s expression. We selected this default number of *cis*-eQTLs based on previous eQTL simulations which select a max of 5 *cis*-eQTLs per gene^7^. Since the number of *trans*-eQTLs may vary significantly across genes in real data^30^, we selected 20 as an intermediate default but simulate between 5 and 50 to identify patterns of EGRET’s performance across a range of values. Causal *cis*-eQTLs were restricted to a 1 Mb window centered at the gene TSS. *trans*-eQTL effects were assigned randomly to variants off-chromosome. To investigate each set of simulation parameters, we randomly selected 100 genes distributed across the 22 autosomes. Genome-wide models of gene expression were built using the Matrix eQTL component of EGRET and exclusively *cis*-eQTL models were generated with FUSION (**Methods**). As described in the previous section, we extracted the maximum CV R^2^ obtained from the set of multivariate regression frameworks in order to compare performance across groups of genes.

Since the genetic architecture of gene expression regulation is a function of both eQTL effect sizes and sparsity, we tested EGRET’s performance at capturing gene expression heritability across different regulatory scenarios. First, to evaluate the effect of different proportions of genome-wide heritability (*h^2^_gw_*) attributed to *trans* regions (*h^2^_trans_*/*h^2^_gw_*), we simulated varying *cis-* and *trans*-eQTL heritabilities that contribute to the *h^2^_gw_* at percentages from 0% to 100% in increments of 10% (**Figure 2A**, **Supplementary Table 1**). The solid lines indicate the proportion of variance explained (R^2^) when different levels (dashed lines) of *h^2^_gw_* are simulated, with *h^2^_gw_* concentrated more in the *cis* region (i.e., low values of *h^2^_trans_/h^2^_gw_*) and more in the *trans* region (i.e., high values of *h^2^_trans_/h^2^_gw_*). As expected, when heritability is more highly concentrated in the *cis* region, Matrix eQTL-only EGRET models capture more of the total heritability as the *cis*-eQTL effects are larger, less dispersed, and thus easier to identify. We also find that Matrix eQTL-only EGRET captures more than half of the genome-wide heritability when it is high (*h^2^_gw_* = 0.6), regardless of the proportion of *h^2^_trans_*. Since previous studies have estimated that the average *cis*-heritability of gene expression is approximately 10%^4,20^ and that *trans*-heritability is approximately two-thirds of total heritability^9,10^, the results for *h^2^_gw_* = 0.3 may be the most representative for the average gene. In this regime (e.g., 70% *h^2^_gw_* in *trans* and *h^2^_gw_* = 0.3), Matrix eQTL-only EGRET captures one-third of the total heritability. While these simulations serve as a baseline, we anticipate that the addition of *trans*-PCO and GBAT in the empirical implementation of EGRET will lead to further capture of the total gene expression heritability and is thus likely to be valuable in downstream applications such as TWAS.

**Figure 2.**
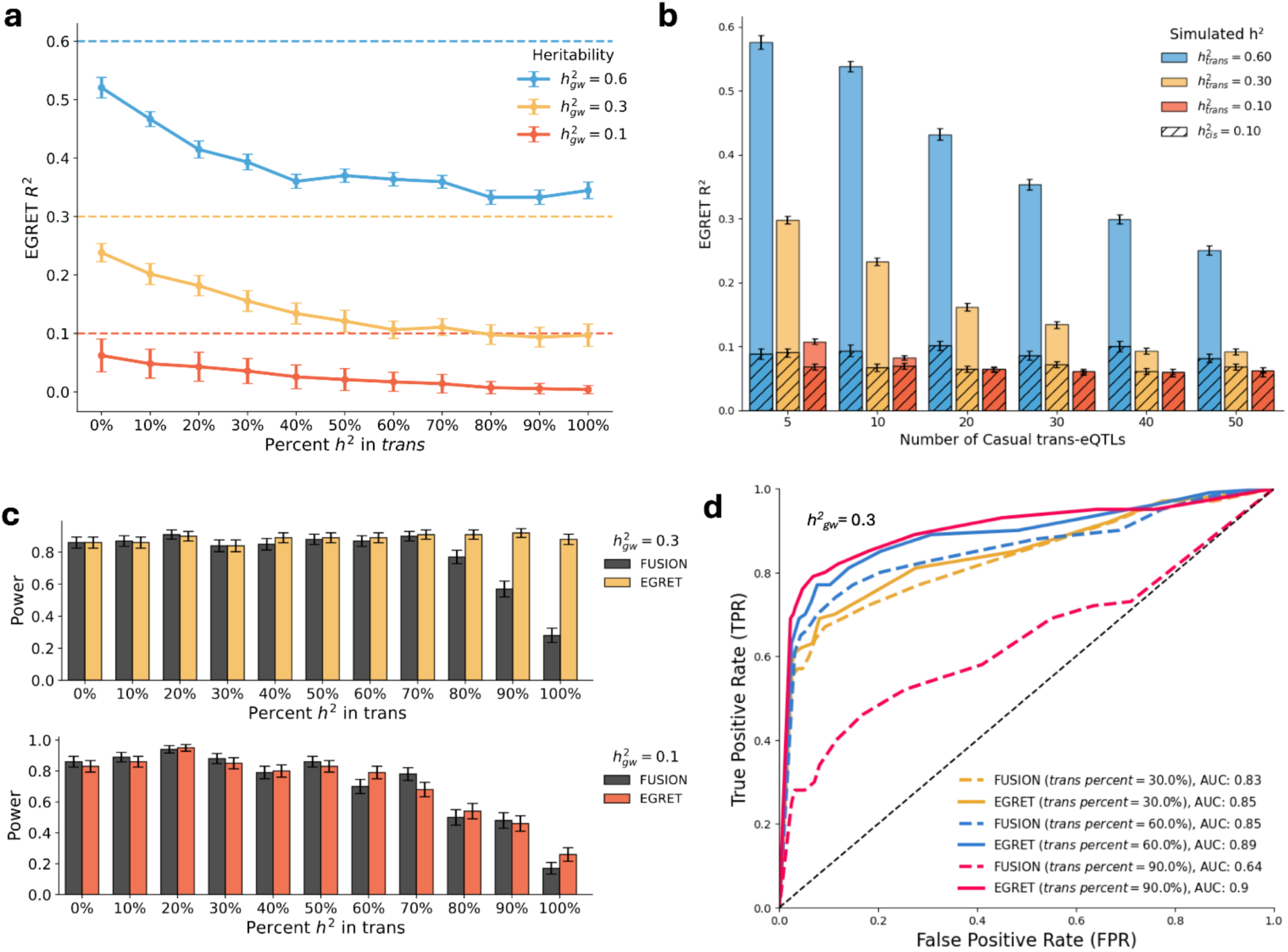
Simulations of genome-wide gene expression models and TWAS. (a) Average Matrix eQTL-only EGRET CV R^2^ at different proportions of total genome-wide heritability in *trans* when there are 20 *trans*-eQTLs. Colors represent different simulated genome-wide heritability levels. Zero percent indicates that all gene expression heritability originates from the *cis* region. Dashed lines represent simulated genome-wide heritability (*h^2^_gw_*) values. (b) Average Matrix eQTL-only EGRET CV R^2^ across specified numbers of causal *trans*-eQTLs simulated. Solid bars represent the genome-wide R^2^. Hashed regions show the amount of variance explained by *cis* variants with non-zero Matrix eQTL-only EGRET effect sizes. Colors indicate *trans*-heritability levels. *cis-*heritability is held constant at 0.1. (c) Power of EGRET-TWAS (yellow, orange) and *cis*-eQTL FUSION-TWAS (gray) to identify causal genes in simulations. (d) Area under the receiver operating characteristic (AUROC) curve shown for EGRET-TWAS (solid lines) and FUSION-TWAS (dashed lines). Colors indicate different percentages of heritability in *trans*. Genome-wide heritability was set to 0.3. In all panels, error bars represent standard errors across 100 independent simulations. All numerical results can be found in **Supplementary Table 1**. Abbreviations: gw, genome-wide; AUC, area under the receiver operating characteristic curve.

Next, we tested the effect of increasing the number of causal *trans*-eQTLs by simulating between 5 and 50 *trans*-eQTLs while keeping the *cis*-to-*trans* heritability ratio constant at several different heritability ratios (**Figure 2B**, **Supplementary Table 1**). Since sampled eQTL effect size magnitudes shrink with increasing numbers of causal variants for a given *cis*-heritability (**Methods**), it is challenging to recover *h^2^_gw_* when the *trans*-eQTL architecture is increasingly polygenic. When *h^2^_trans_*/*h^2^_gw_* is large (i.e., *h^2^_gw_* = 0.6), we recover about half of *h^2^_trans_* at the largest tested number of *trans*-eQTLs. However, when the ratio is small (i.e., *h^2^_gw_* = 0.1), beyond 10 *trans*-eQTLs, the Matrix eQTL-only EGRET model does not identify any SNPs that meaningfully contribute to explaining the variance of gene expression. Therefore, in real data, our method should be most advantageous for genes with larger proportions of *h^2^_trans_*, seemingly regardless of the number of *trans*-eQTLs, and still modestly advantageous for genes with smaller proportions of *trans*-heritability but fewer total *trans*-eQTLs. The hashed boxes represent the variance explained by predictive SNPs (non-zero estimated effect sizes) in the *cis* region and align with the variance explained by standard FUSION models. As expected, eQTL cohort size also strongly correlates with Matrix eQTL-only EGRET’s ability to create accurate gene expression prediction models (**Supplementary Figure 2**). We again note that in real data analysis, we expect EGRET to perform better than in this simplified simulation framework, since we utilize functional information such as co-expression patterns that should help reveal *trans*-regulatory loci.

To evaluate the second point of assessing how well genome-wide gene expression models comparatively perform at nominating disease-critical genes in TWAS, we compared the relative ability of Matrix eQTL-only EGRET and FUSION^4^ gene models to detect genes designated as disease causal in our simulations. We refer to our framework as EGRET-TWAS and the standard FUSION framework as FUSION-TWAS. We simulated polygenic diseases and their corresponding GWAS summary statistics based on gene expression and environmental noise (**Methods**). In all simulations, 10 of the 100 genes simulated are causal and their eQTLs mediate 10% of total disease heritability, based on previously published empirical results^32–34^. Polygenic diseases were generated for all individuals based on the previously defined eQTL effect sizes and the predetermined contribution of gene expression to the polygenic disease. Corresponding GWAS summary statistics were produced using a linear model including 5 genotype PCs as covariates. To compute TWAS *z*-scores, a weighted genome-wide burden test was performed using the EGRET and FUSION models (**Methods**). Finally, we calculated power and FPR across 100 independent simulations representing each of various simulation parameters. Sensitivity and specificity were simultaneously evaluated using the area under the receiver operating characteristic (AUROC) curve; we used 15 evenly spaced p-value thresholds ranging from 0 to 1.

In these analyses, we anticipated that modeling the *trans*-component of gene expression would allow for greater colocalization with GWAS loci and thus substantially increase power to identify disease-critical genes in TWAS. While both methods exhibit comparable power at relatively low genome-wide heritability (*h^2^_gw_* = 0.1), EGRET-TWAS demonstrates improved power at larger heritabilities (i.e., *h^2^_gw_* > 0.1), specifically for genes with > 70% of h^2^_gw_ in *trans* regions (**Figure 2C**). Notably, this critical threshold is representative of realistic gene expression architecture, according to previous studies that estimated approximately two-thirds of gene expression heritability resides in *trans* regions^9,13^. We observe a very similar pattern for *h^2^_gw_* = 0.6 and *h^2^_gw_* = 0.9 (**Supplementary Figure 3**). While **Figure 2B** indicates that EGRET can provide gains in explaining the variance of gene expression relative to standard FUSION (approximated by the hashed bars), **Figure 2C** suggests that these gains translate to increased TWAS power for genes with more than 70% of h^2^_gw_ explained by *trans*-variants and *h^2^_gw_* > 0.1 (**Supplementary Table 1**). Under identical simulation settings, EGRET-TWAS maintains FPRs comparable to FUSION-TWAS (**Supplementary Figure 4**). We note that the lower FPR of FUSION-TWAS at high proportions of *trans-*heritability may be explained by its relatively lower power, as it only models *cis*-eQTLs. We also investigate the effect of varying the number of *trans*-eQTLs on TWAS power and FPR within EGRET’s framework and observe no significant changes across our simulated range (**Supplementary Figure 5**). Additionally, comparisons of AUROC show that EGRET-TWAS consistently outperforms standard FUSION-TWAS; this advantage becomes more pronounced at increasing proportions of *trans-*heritability (**Figure 2D**, **Supplementary Table 1**). Together, these simulations highlight that by modeling and incorporating putative *trans*-eQTL effects, EGRET enhances gene expression prediction models, which may further aid in identifying disease-critical genes in TWAS.

### Evaluating EGRET gene model performance across tissues and competing methods

We next set out to understand how EGRET could augment our understanding of genetic regulation of gene expression across the transcriptome of different human tissues. To this end, we applied EGRET to paired RNA-sequencing (RNA-seq) and genotype data from 49 tissues assayed by the GTEx Consortium^7^, with sample sizes ranging from 65 in kidney cortex to 605 in skeletal muscle. Standard covariates, including sex, age, sequencing batch, 5 genotype PCs, and between 15 and 60 Probabilistic Estimation of Expression Residuals (PEER) factors^35^, previously determined by sample size^25^, were regressed out in all analyses (**Methods**). For Weighted Gene Co-expression Network Analysis (WGCNA)-based clustering used in *trans*-PCO, we applied a relatively reduced set of covariates. Specifically, we regressed out only the top 10 PEER factors, based on the rationale that higher-order PEER factors (e.g., those that explain less variance) may capture *trans*-acting regulatory variation. This approach is consistent with the original *trans*-PCO and GBAT frameworks^17,19^ and other studies that restrict expression PC correction^36^. Notably, these top 10 PEER factors correlate strongly with GTEx death code (**Supplementary Figure 6**).

We ran each *trans*-eQTL detection method (Matrix eQTL, *trans*-PCO, and GBAT) independently within each of five CV folds to avoid overfitting. Each method was adapted for EGRET’s application (**Methods**). While the genetic architecture of a gene can be described by the relative proportion of *cis*- versus *trans*-eQTL regulation, we calculated the CV R^2^ (i.e., using predictions on the held-out fold) for each of three sets of variants: *cis-*only, *trans-*only, and all variants genome-wide. For variance explained by *cis*-eQTLs, we use a conservative 5 Mb region centered on the gene TSS to maximally include tagging effects of *cis*-eQTLs beyond the 1 Mb *cis* window and avoid double-counting decaying *cis*-eQTL effects into our calculation of explained variance attributed to *trans*-eQTLs. For variance explained by putative *trans*-eQTLs, we used all predictive features beyond the 5 Mb region. For variance explained by all putative eQTLs genome-wide, we used all predictive features with no exclusions; only for this analysis, we let EGRET models default to the FUSION model when EGRET fails to produce a model (R^2^ < 0).

Across 49 tissues, EGRET produced 353,408 predictive gene expression models (R^2^ > 0, p < 0.01). We observed the average R^2^ of EGRET models is comparable to FUSION *cis*-variant models (**Figure 3A** left, FUSION R^2^ = 0.049, Standard Error (SE) = 0.002, EGRET R^2^ = 0.050, SE = 0.002, **Supplementary Table 2**). For 28 tissues, EGRET models exhibit a significant improvement over FUSION models (one-sided Wilcoxon rank-sum test p < 0.005) (**Figure 3A** left). Considering genes with successful EGRET models (R^2^ > 0, p < 0.01) across each tissue and each regularization strategy from the EGRET framework, xtune LASSO most frequently explained the largest variance of gene expression across larger sample size tissues, while elastic net and standard LASSO performed better for smaller sample size tissues (**Supplementary Figure 7**). Notably, we found that EGRET generates predictive models for an average of 1,355 genes per tissue that did not have a significant model (R^2^ p < 0.01) with FUSION (**Figure 3A**, right). In other words, *cis*-SNPs did not explain a significantly nonzero proportion of expression variance for these genes (FUSION R^2^ p > 0.01) and thus could not have been analyzed for disease colocalization with TWAS. There were also approximately 1,240 genes per tissue for which *only* the standard *cis*-eQTL model resulted in a significantly nonzero R^2^; this may be the result of EGRET adding non-informative features to the models, resulting in weaker performance in genome-wide prediction tasks.

**Figure 3.**
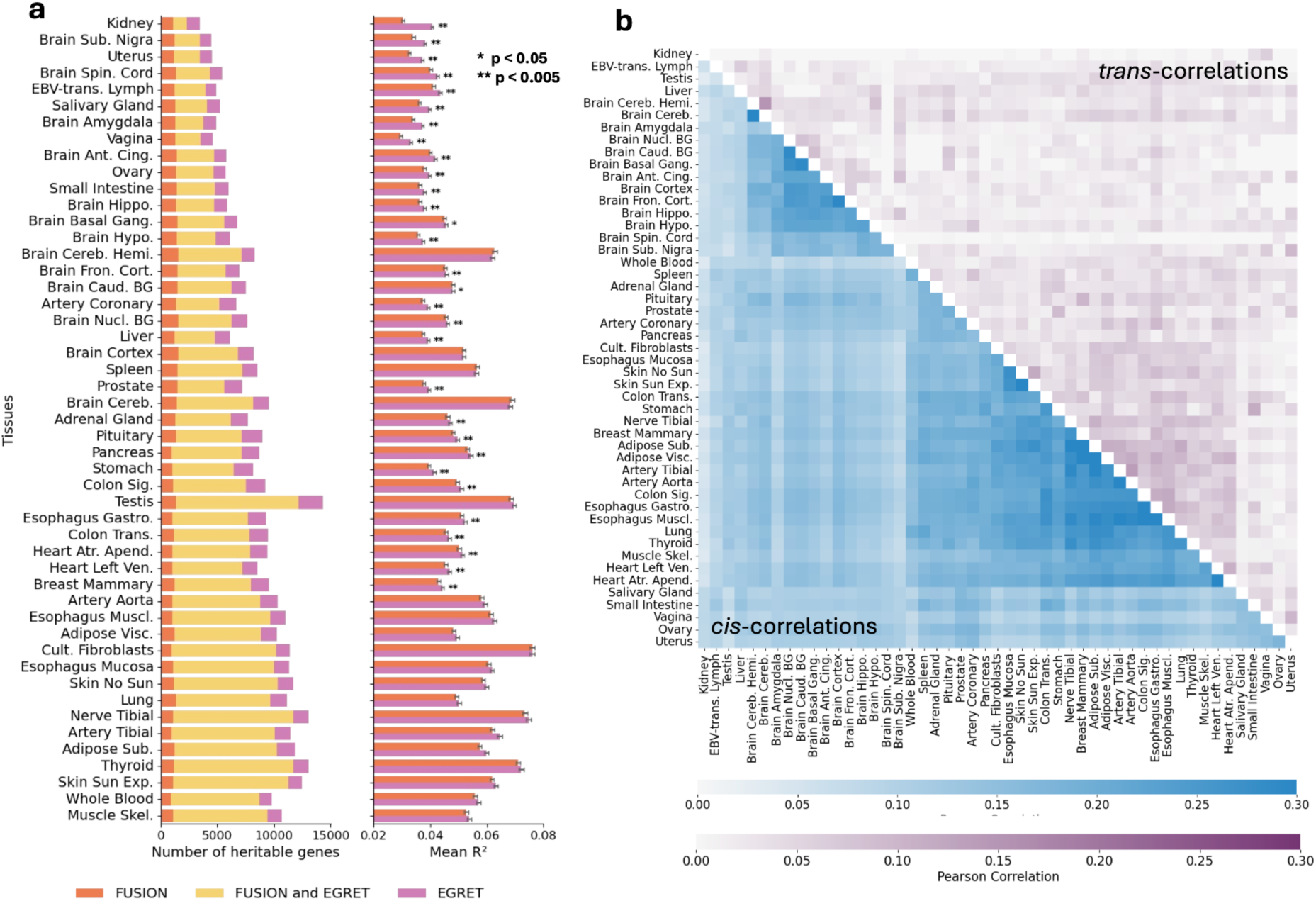
Benchmarking EGRET gene expression prediction models with *cis*-eQTL FUSION models across tissues. (a) (Left) Number of genes across tissues with a significantly nonzero amount of gene expression explained by *cis*-variants only (pink), EGRET models only (purple), or both simultaneously (yellow) (CV R^2^ > 0, p < 0.01). Tissues are ranked by smallest to largest sample size from top to bottom. (Right) Average model performance (CV R^2^) across tissues using FUSION compared to EGRET models. (b) Average pairwise Pearson correlation of gene expression prediction profiles across tissues after imputing expression into individuals from the 1000 Genomes Project. For the lower triangle, we exclusively use weighted *cis*-variants from EGRET models, whereas for the upper triangle, we exclusively use weighted *trans*-variants from EGRET models. Tissues are ordered according to hierarchical clustering of *cis*-correlations. All numerical results can be found in **Supplementary Table 2**. Abbreviations: Visc, Visceral; Spin, Spinal; Sub., Substantia; Ant. Cing., Anterior Cingulate; Gang. Ganglia; Hippo, Hippocampus; Cereb. Hemi., Cerebellar Hemisphere; Fron. Cort., Frontal Cortex; Caud. BG, Caudate Basal Ganglia; Nucl. BG, Nucleus Accumbens Basal Ganglia; Cereb., Cerebellum; Sig., Sigmoid; Trans., Transverse; Ven., Ventricle; Muscl., Muscularis; Cult., Cultured; Sub., Subcutaneous; Exp., Exposed; Skel., Skeletal.

Next, we investigated the perceived sharing of *cis-* and *trans*-effects across tissues. Within the GTEx consortium, it was observed that only 3.8% of *trans*-eQTLs were shared across 3 or more tissues compared to 25.3% of *cis*-eQTLs^37^. Therefore, we hoped that we could identify greater sharing of *trans*-eQTL effects across tissues by leveraging EGRET models which overcome some limitations of univariate SNP-gene testing. Thorough investigation into tissue co-regulation has previously been conducted between *cis*-eQTLs^25,34,37^, however, few have investigated *trans*-genetic co-regulation^25^. Here, we use expression prediction models learned by EGRET to assess the differences in gene expression co-regulation driven by *cis-* and *trans-*regulatory variants (**Figure 3B**, **Supplementary Table 2**). First, we recognize that calculating genetic correlations is not straightforward when effect sizes are estimated with sparse regularization models. To overcome this, we instead compute correlations of imputed expression by taking the product of EGRET model weights (separated into *cis-* and *trans-*SNPs (> 5 Mb)) and the standardized genotypes of 489 European-ancestry individuals from the 1000 Genomes Project^38^. We then calculated the pairwise correlation of these *cis*- or *trans*-predicted expression values for a given gene across different pairs of tissues. For a given pair of tissues, we report the average of these Pearson correlations across the set of genes with a significantly predictive model (R^2^ > 0, p < 0.01) in at least one of the tissues. Exemplar pairs of tissues with the highest correlation of *cis*-eQTL predictions were (1) brain cerebellum and the cerebellar hemisphere (*ρ* = 0.35, SE = 6e-5) and (2) tibial artery and aorta artery (*ρ* = 0.34 SE = 4e-5); reassuringly, these pairs of tissues also have higher correlations in their *trans*-predictions (*ρ* = 0.15, SE = 0.005 and *ρ* = 0.14, SE = 0.002, respectively) and relative to other tissue pairs. However, on average, the pairwise correlation of *cis*-predictions between tissues was 0.146 (SE = 2.37e-05), compared to 0.030 (SE = 1.06e-05) for *trans*-predictions. While this result can be explained by a lack of power to identify *trans*-regulatory loci for a given gene, it may also indicate that these loci are more tissue-specific than *cis*-eQTLs. On the contrary, some tissue pairs had relatively high correlations in their *trans*-predictions, e.g., stomach and small intestine (*ρ* = 0.12, SE = 0.007), pituitary gland and breast tissue (*ρ* = 0.10, SE = 0.004), and brain substantia nigra and the uterus (*ρ* = 0.097, SE = 0.03). These results are consistent with cell-cell signaling cascades involving hormones and lineage-determining transcription factors known to involve these very pairs of tissues^39–42^. Ultimately, hierarchical clustering of *trans*-correlations revealed fewer consensus groups of similar tissues (**Supplementary Figure 8**), while clustering of *cis*-correlations revealed two predominant groups–brain and non-brain/non-reproductive tissues–consistent with previous correlations of *cis*-eQTL effect sizes^25^. Sample size is an important consideration for this analysis, as low sample size tissues are less likely to observe strong *cis*- or *trans*-correlations, due to higher variance in their effect size estimates. Overall, we identified a shared underlying genetic basis for several dynamic signaling processes that bridge different anatomical regions of the human body, which could not be identified using *cis*-eQTL associations alone.

The detection of disease-critical genes via TWAS relies on accurately modeling the genetic component of gene expression. Therefore, we sought to evaluate the amount of gene expression variance EGRET models capture compared to other genome-wide methods such as MOSTWAS and BGW-TWAS^21,22^. MOSTWAS utilizes mediator-enriched TWAS (MeTWAS) and distal-eQTL prioritization via mediation analysis (DePMA) to nominate distal regulatory variants and improve gene expression models. On the other hand, BGW-TWAS iteratively selects variants using Bayesian variable selection regression to build genome-wide models. However, as discussed earlier, cross-mappability, or the incorrect mapping of sequencing reads to similar genomic regions, introduces the potential for false positive associations across these methods, including our own. While we previously discussed leveraging mappability scores to reduce bias during the EGRET model building stage, in this analysis, we restricted our comparisons to genes that are not cross-mappable (i.e., have high mappability scores^23^, **Methods**). For a subset of 4,854 genes with a cross-mappability score greater than 0.99 in whole blood, we computed the average prediction R^2^ across genes for each method. For genes which did not converge, the R^2^ for that gene was set to 0. We elected to perform this benchmarking analysis in whole blood due to the availability of independent whole blood eQTL datasets for validation, such as the Depression Gene Networks (DGN) cohort^43^ (analyzed in the next section). MOSTWAS and BGW-TWAS had nearly equivalent mean R^2^ compared to EGRET across these genes (one-sided paired t-test p > 0.05) (EGRET R^2^ = 0.047, SE = 0.0014; MOSTWAS R^2^ = 0.046, SE = 0.0014; BGW-TWAS R^2^ = 0.047, SE = 0.0014) (**Supplementary Figure 9**).

### Interpreting functional propensity of trans-regulatory loci identified by EGRET models

Each constituent *trans*-eQTL mapping method within EGRET is powered to identify different types of variants: Matrix eQTL identifies those with the largest effect sizes, GBAT identifies those that are *cis*-eQTLs of upstream or downstream genes, and *trans*-PCO identifies those with most consistent regulation across co-expressed genes. To this end, we sought to assess the independent contribution of each *trans*-eQTL mapping method to EGRET by reanalyzing the GTEx tissue data and applying each method in isolation to construct gene expression prediction models. This decomposition allowed us to evaluate the contribution of each mapping method to the performance of EGRET. For this analysis, we restrict to genes whose expression variance can be substantively explained by *trans*-variants alone, defined as significantly nonzero CV R^2^_trans_ in cross-validation (p < 0.01); this refers to using all variants beyond the 5 Mb window around the gene TSS and their coefficients that were fit by EGRET in the previous section. We refer to these genes as *trans*-heritable. Across 49 GTEx tissues, we identified 12,317 gene-tissue pairs with a significantly nonzero *trans*-heritable component (R^2^_trans_ p < 0.01). For these genes, EGRET models explained 33.3% more variance than FUSION models (EGRET average R^2^ = 0.104, FUSION average R^2^ = 0.078). Then, to control for method-specific differences in modeling *cis*-eQTL features, we included all *cis*-variants and method-specific *trans*-variants as variables in the multivariate regressions. Finally, we calculated the method-specific CV R^2^_trans_ using only variants beyond the 5 Mb window and the method-specific CV R^2^ which includes all *cis*-variants and *trans*-variants specifically nominated by each method. We use the method-specific R^2^_trans_ to determine which method was most informative for a given gene in the *trans* region and we use the method-specific R^2^ to calculate improvements from the *cis*-eQTL FUSION model.

Averaged across tissues, Matrix eQTL was 2.1x more likely to produce the most predictive model relative to GBAT, which in turn was 3.6x more likely to produce the most predictive model compared to *trans*-PCO (**Figure 4A**, left, **Supplementary Table 3**). For this comparison, we selected 10 biologically independent and anatomically representative tissues of relatively large sample size.

**Figure 4.**
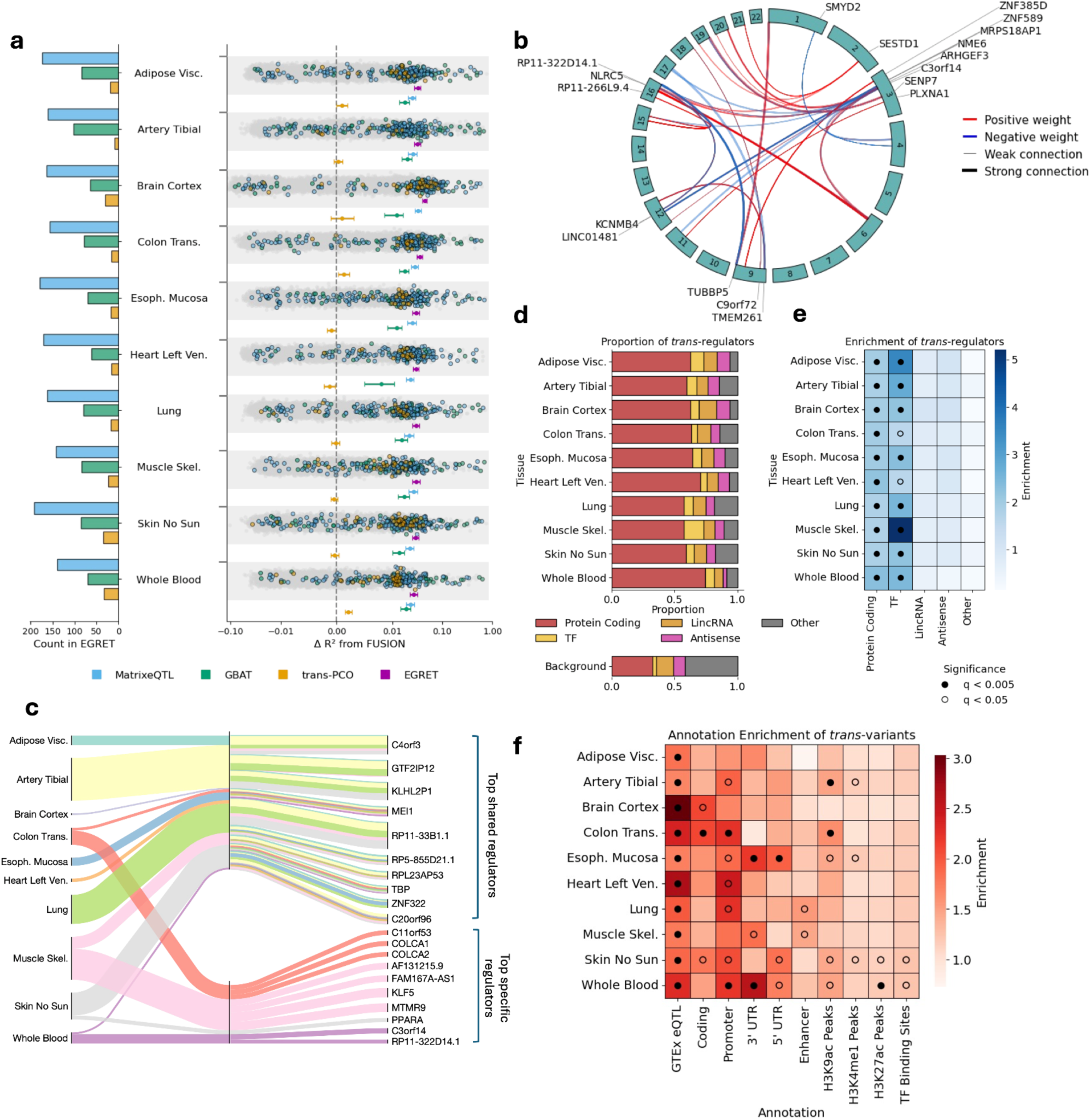
EGRET gene models improve gene expression prediction by modeling known *trans*-regulators. (a) (Left) Frequency with which each *trans*-eQTL detection method produces the largest method-specific R^2^_trans_ across *trans*-heritable EGRET genes. (Right) Change in R^2^ across method-specific models and EGRET relative to *cis*-variant FUSION models, with bars representing +/- standard error around the mean. Gene-level changes for *trans*-heritable EGRET genes are colored by the method producing the largest method-specific R^2^_trans_, while non-*trans*-heritable genes are shown in gray. (b) Plot of top *trans*-regulators (>5 downstream targets) for GTEx whole blood EGRET models. (c) Top tissue-shared and tissue-specific *trans*-regulators based on the number of downstream targets. (d) Proportion of tissue-specific *trans*-regulator genes in gene classes. “Other” includes pseudogenes and other non-coding RNA gene products (e) Enrichment of *trans*-regulator genes in gene classes. (f) Enrichment of weighted *trans*-variants in functional annotations. All numerical results can be found in **Supplementary Tables 3-5.** Abbreviations: Visc, Visceral; Trans., Transverse; Ven., Ventricle; Skel., Skeletal.

In **Figure 4A** (right), we quantified the change in prediction accuracy (ΔR^2^) for the EGRET and method-specific models relative to FUSION across those 12,317 *trans*-heritable genes. While *cis*-eQTL effects are included in each model, we attribute any differences in performance to differences in the *trans*-variants prioritized by each method. Among the constituent methods, Matrix eQTL (blue bars) and GBAT (green bars) models generated larger R^2^ values than *trans*-PCO (yellow bars). In fact, we found that using *trans*-PCO in isolation often produced gene models with nearly the same performance as FUSION (average ΔR^2^ = 2e-4, SE = 3e-4), which may indicate that the prioritized *trans*-variants add more noise than predictive features to the model, detracting from fitting appropriate weights for *cis*-variants. This may be expected as *trans*-PCO does not explicitly identify which genes within a module are associated with particular variants. For the ΔR^2^ attributed to EGRET, we further show the gene-level differences in prediction and we color each gene by the constituent method that resulted in the largest method-specific R^2^_trans_. The average increase in prediction due to EGRET across all *trans*-heritable genes across these 10 tissues was ΔR^2^ = 0.032 (SE = 0.0045). For genes that were not *trans*-heritable, we observe some instances of striking improvement with EGRET models (right-most gray dots), suggesting that variants within the 1 Mb and 5 Mb windows are substantially boosting prediction while variants beyond 5 Mb do not. Most notably, our results highlight the benefit of leveraging orthogonal *trans*-eQTL detection approaches capturing mechanistically distinct features, as the integrative EGRET model consistently outperforms each method in isolation. For example, EGRET improves the prediction of *HBG2* in whole blood from R^2^ = 0.18 (p = 8.3e-27) to R^2^ = 0.34 (p = 1.42e-53), representing an 89% increase in the variance explained. This gene has previously been reported to be regulated by several *trans*-eQTLs and is implicated in beta-thalassemia, a blood disorder^44^. Markedly, several genes that did not have a predictive FUSION are rather strongly regulated by *trans* loci. For example, *CLEC18B* (EGRET R^2^ = 0.32, p = 9.26e-52) is strongly regulated by *cis*-SNPs of several transcription factors such as *ZNF683* and *PBX1*; FUSION models for this gene failed to converge. Other genes which show very striking *trans*-regulation are *NPIPB15* (EGRET R^2^ = 0.54, p = 3.77e-99; FUSION R^2^ = 0, p = 1), *WASHC2A* (EGRET R^2^ = 0.33, p = 2.11e-51; FUSION R^2^ = 0.20, p = 5.28e-30). We have released a full list of genes and their performance metrics on Zenodo (**Data Availability**).

Next, we investigated the potential functional mechanisms of prioritized *trans*-variants in EGRET models, e.g., variants that were assigned a nonzero coefficient or weight. Since *cis*-eQTLs of upstream regulatory genes are hypothesized to explain a large fraction of the mechanisms by which *trans*-eQTLs act^45^, we set out to identify potential “*trans*-regulator” genes of *trans*-heritable EGRET genes. To identify potential *trans*-regulators, we first use previously calculated *cis*-eQTL associations^25^ to link weighted *trans*-variants to their *cis*-regulated genes (**Methods**). However, if the weighted *trans*-variant is not a *cis*-eQTL of any gene, we assign the most proximal expressed gene as a potential *trans*-regulator of the gene modeled by EGRET. This assumes that the *trans*-eQTLs are acting as *cis*-regulators of the nearest expressed genes.

We report top *trans*-regulators (those with at least 5 downstream gene targets) of genes expressed in GTEx whole blood tissue (**Figure 4B**, **Supplementary Table 4**); *trans*-regulators for other tissues can be found on our Zenodo (**Data Availability**). We depict the downstream targets of *trans*-regulators via arcs, where the *trans*-regulator gene names are labeled. Among 197 *trans*-heritable genes in whole blood, we identified 636 weighted *trans*-variants, implicating 526 unique potential *trans*-regulators. Many of these *trans*-regulators are known transcription factor genes such as *ZNF589*, critical for cell viability in the hematopoietic system^454^. Notably, most *trans*-regulators had one downstream target yet *NLRC5* had 17 connections, representing a major regulator of MHC class I gene^47^. Others such as *KCNMB4* and *PLXNA1* are receptor proteins, suggesting that *trans*-regulation may involve cell-cell signaling pathway^48,49^. Finally, we highlight *LINC01481*, a long non-coding RNA found to be upregulated during immune cell invasion within ischemic stroke patients and highlighted as a key locus in osteoarthritis, which is driven by myeloid-derived osteoclasts^50,51^. Altogether, our findings highlight diverse mechanisms that result in *trans*-regulation of other genes.

Intriguingly, we observe many of the same *trans*-regulators acting in other tissues and provide a multi-tissue overview of our results in **Figure 4C** (**Supplementary Table 5**). We then assessed the tissue-specificity of the downstream targets of these *trans*-regulators. We refer to tissue-shared regulators as those that appear in at least 4 of the 10 representative tissues, while tissue-specific regulators appear in only a single tissue. Band thickness denotes the relative number of genes within each tissue regulated by that specific *trans*-regulator; numerical values are provided in **Supplementary Table 5**. Notable examples include *C3orf14* in whole blood which is a *trans*-regulator for 17 downstream targets and is known to be downregulated in patients with acute myeloid leukemia suggesting it may have a critical role in hematopoietic cell regulation^52,53^.

Among other tissue-specific *trans*-regulators, we found *PPARA* to be exclusive to skin tissue; while this gene is a pleiotropic regulator, it has specifically been found to be crucial for skin homeostasis^54,55^. Separately, *COLCA1* and *COLCA2*, exclusive to transverse colon tissue, have previously been implicated in driving colon cancer, stressing its important role as a tissue-specific transcriptional co-activator^56^. On the contrary, several tissues exhibited no tissue-specific *trans*-regulators, e.g., visceral adipose, esophageal mucosa, tibial artery, brain cortex, lung, and heart left ventricle, which may be explained by reduced power due to smaller sample sizes and therefore generally shared patterns of the most prominent types of gene regulation relative to one another. We also identified tissue-shared *trans*-regulators that are known to play generic housekeeping roles. For example, *TBP* encodes a TATA-binding protein, a universal transcription factor essential for initiation by all three RNA polymerases, and is found to be a *trans*-regulator in 7 tissues^57^. Surprisingly, some of the most commonly shared *trans*-regulators are non-coding RNAs. For example, *RPL23AP53* is a pseudogene documented to have a role in melanoma and is associated with cellular proliferation^58^. Others such as *RP5-855D21.1*, *RP11-33B1.1*, *GTF2IP12*, and *KLHL2P1* corroborate previous work that demonstrates that pseudogenes play a substantive role in gene expression regulation^59,60^. All identified *trans*-regulators and their target genes across tissues can be found in **Supplementary Table 6**.

Next, we assessed the validity of putative *trans-*regulatory loci identified by EGRET models with functional enrichment analysis. First, we investigated the “*trans*-regulators” in these loci; second, we investigated the variants themselves. We first employed gene class nomenclature from GTEx and a comprehensive list of transcription factors from Lambert and colleagues^609^ (**Methods**). We classified *trans*-regulator genes and, for comparison, the set of all expressed genes in across GTEx as either protein-coding, transcription factor (TF), long-intergenic non-coding RNA (lincRNA), anti-sense RNA, and other non-coding gene products including pseudogenes. While we only constructed EGRET models for protein coding, lincRNAs, and antisense transcripts, we allowed other types of genes (i.e., pseudogenes) to be identified as *trans*-regulators, such as when using GTEx *cis*-eQTL summary statistics to linking weighted *trans*-variants to their *cis*-regulated genes. We then determined the proportion of genes belonging to each category for *trans*-regulators versus the set of all genes (**Figure 4D**).

Consistent with other *trans*-eQTL studies^18,30^, *trans*-regulator genes were significantly enriched for protein coding genes. We also found that 8 tissues showed very significant enrichment for transcription factor genes (FDR < 0.005) (**Figure 4E**). Second, to assess the regulatory elements harboring weighted *trans*-variants, we calculated their enrichment across various functional categories including GTEx *cis*-eQTLs^25^ and annotations from the LD score regression (LDSC) framework^62^. Enrichment was calculated as the number of variants falling within annotated regions divided by the expected number to fall within these regions based on the total number of weighted *trans*-variants observed; we use a hypergeometric distribution to calculate the corresponding enrichment p-value, as done previously^18^ (**Methods**). Overall, we observed substantial enrichment of weighted *trans*-variants in regions of the genome associated with transcriptional regulation, although patterns tended to be tissue-specific (**Figure 4F**, **Supplementary Table 5**). For example, weighted *trans*-variants across all 10 representative tissues exhibited significant enrichment of GTEx *cis*-eQTLs (cross-tissue average *-log_10_*(p) = 22.03). Our analysis revealed strong enrichments in the untranslated flanking regions of genes (3’ UTR and 5’ UTR) in esophageal, skeletal, skin, and blood tissues. Interestingly, we also observed an enrichment in coding regions for four tissues, most likely representing synonymous mutations that are unlikely to alter protein structure. In the epigenetic elements H3K9ac, H3K4me1, and H3K27ac, which mark active enhancers and promoters^63^, *trans*-variants were enriched with greater tissue-specificity. Similarly, enhancer elements identified by chromatin accessibility assays also produced tissue-specific enrichments in lung and muscle tissue.

In order to replicate our results beyond the GTEx cohort, we used the Depression Gene Networks (DGN) dataset which includes whole blood RNA-seq and genotype data from 922 European individuals^43^. First, we compared EGRET and FUSION models to predict expression in the DGN cohort (**Methods**). For this analysis, we considered 6,278 genes with a significantly nonzero EGRET R^2^ (p < 0.01) in GTEx whole blood and 6,620 genes in the same tissue with a significantly nonzero FUSION R^2^ (p < 0.01). On average, FUSION and EGRET performed similarly across these sets of genes (FUSION R^2^ = 0.113, SE = 0.002; EGRET R^2^ = 0.109, SE = 0.002). Second, we hypothesized that EGRET may outperform FUSION in a subset of genes with strong *trans*-heritability. If so, this could indicate that EGRET is identifying replicable *trans*-regulatory loci. Therefore, in a subset of genes with significantly nonzero R^2^_trans_ in EGRET, we investigated the predictive performance of FUSION and EGRET models. We varied the p-value threshold used to characterize a gene as having significantly nonzero R^2^_trans_ (default p < 0.01). For more lenient thresholds, FUSION and EGRET result in similar R^2^ across the corresponding gene subsets (**Supplementary Figure 10**), but at p-values smaller than 1e-6, EGRET model performance begins to overtake FUSION model performance. This suggests that *trans*-regulatory loci identified by EGRET in the most highly significant *trans*-heritable genes are reproducibly explaining more variance than *cis*-eQTLs in an independent eQTL cohort. We also assessed MOSTWAS and BGW-TWAS models in the DGN cohort; these models perform similarly to EGRET across the same set of non-cross-mappable genes used to benchmark performance in GTEx (EGRET R^2^ = 0.032, SE = 0.001), (MOSTWAS R^2^ = 0.034, SE = 0.001), and (BGW-TWAS R^2^ = 0.031, SE = 0.001) (**Supplementary Figure 11**).

### EGRET identifies genes that may confer disease risk via trans-regulatory regions

Modeling genome-wide regulatory effects on gene expression not only has the potential to improve gene expression prediction model performance, but also to identify new disease-critical genes that harbor disease risk specifically in their *trans*-genetic components. As conventional TWAS methods utilize *cis*-variant prediction models, such as FUSION^4^ and PrediXcan^20^, some disease-critical regulatory mechanisms may be excluded from consideration leading to false negatives. Therefore, we augment the traditional summary statistics-based *cis*-eQTL TWAS framework to include distal variant effects (**Methods**). We then apply EGRET models trained in 49 GTEx tissue models to perform a TWAS across 78 complex traits and diseases (average *N* = 302K; **Methods**). We identified 1,351,983 gene-tissue-disease associations at FDR < 5% across 24,190,893 gene-tissue-disease trios tested using EGRET-TWAS. A full list of gene-tissue-disease associations can be found on Zenodo (**Data Availability**). Of these significantly associated gene-tissue-disease trios, 450,825 (or 33%) were not detected by FUSION. 43% of the 450,825 associations did not have a predictive FUSION model (e.g., R^2^ was not significantly nonzero), whereas the other 57% obtained a predictive FUSION model but subsequently were not significantly associated via TWAS (FDR > 5%), indicating that EGRET-TWAS nominates a substantial set of associations missed by *cis*-eQTL approaches. Moreover, the preponderance of gene-tissue pairs without a predictive FUSION model indicates that their expression and disease associations are largely driven by *trans*-regulatory mechanisms rather than *cis*-genetic effects. Consistent with that, 23,398 gene-tissue-disease associations are the result of a *trans*-heritable gene (R²*_trans_* > 0, p < 0.01), suggesting that *trans*-eQTL effects play an important role in uncovering disease-associated genes.

FUSION identified 1,713,834 gene-tissue-disease associations at FDR < 5%. While the majority of TWAS associations are shared between FUSION and EGRET, 812,676 of these 1,713,834 (or 47%) associations were not detected by EGRET. We attribute this discrepancy largely to two factors. First, EGRET’s genome-wide expression models incorporate a larger set of variants, which can reduce predictive performance for some genes due to increased model complexity and noise. Second, the inclusion of *trans*-variants can introduce effects with opposing directions on disease risk, attenuating associations that are detected by *cis*-only models. Despite these differences, we observe strong concordance in TWAS *z*-scores between FUSION and EGRET across a subset of tissues and traits (Pearson R = 0.87, p < 1e-308 **(Supplementary Figure 12**), indicating that the overall genetic signals captured by the two methods remain highly consistent.

We also benchmarked EGRET-TWAS against BGW-TWAS and MOSTWAS. Across the same subset of 4,854 non-cross-mappable genes in whole blood, we identified 10,182 gene-tissue-disease associations at 5% FDR with EGRET-TWAS, 7,799 associations with BGW-TWAS and 9,820 with MOSTWAS. Of those genes which are identified by EGRET, 2,900 associations were not identified by MOSTWAS and 5,498 associations were not identified by BGW-TWAS. Overall, we found significant correlation of TWAS *z*-scores across all traits in whole blood tissue (R = 0.58, p < 1e-308 for EGRET with BGW-TWAS and R = 0.81, p < 1e-308 for EGRET with MOSTWAS) (**Supplementary Figure 13**).

We use whole blood tissue to exemplify genome-wide EGRET-TWAS *z*-scores for 15 blood and immune related traits (**Figure 5A**, **Supplementary Table 7**). Significant EGRET gene associations which were not FDR significant using FUSION models and are *trans*-heritable are emphasized with a black circle. First, we discuss exemplar associations in which EGRET recapitulates previous findings of FUSION models, providing reassurance that EGRET models are capable of capturing disease-critical *cis*-genetic regulation as well. Of the shared associations (at 5% FDR) among FUSION and EGRET models, several show increased significance in EGRET models. One such case is *ALDH2* (EGRET-TWAS p = 6.88e-51, FUSION-TWAS p = 1.13e-42) for diastolic blood pressure in whole blood. While this gene has been previously implicated for blood pressure^64^, EGRET models show increased significance likely due to its *trans*-genetic component (FUSION R^2^=0.044, EGRET R^2^=0.090), suggesting that disease-critical effects colocalize with both the *cis*-genetic and *trans*-genetic variants of this gene. Notably, two of the *trans*-regulators (defined earlier) of *ALDH2, MCTP2* and *MIR4458HG,* have previously been associated with cardiovascular events^65,66^.

**Figure 5.**
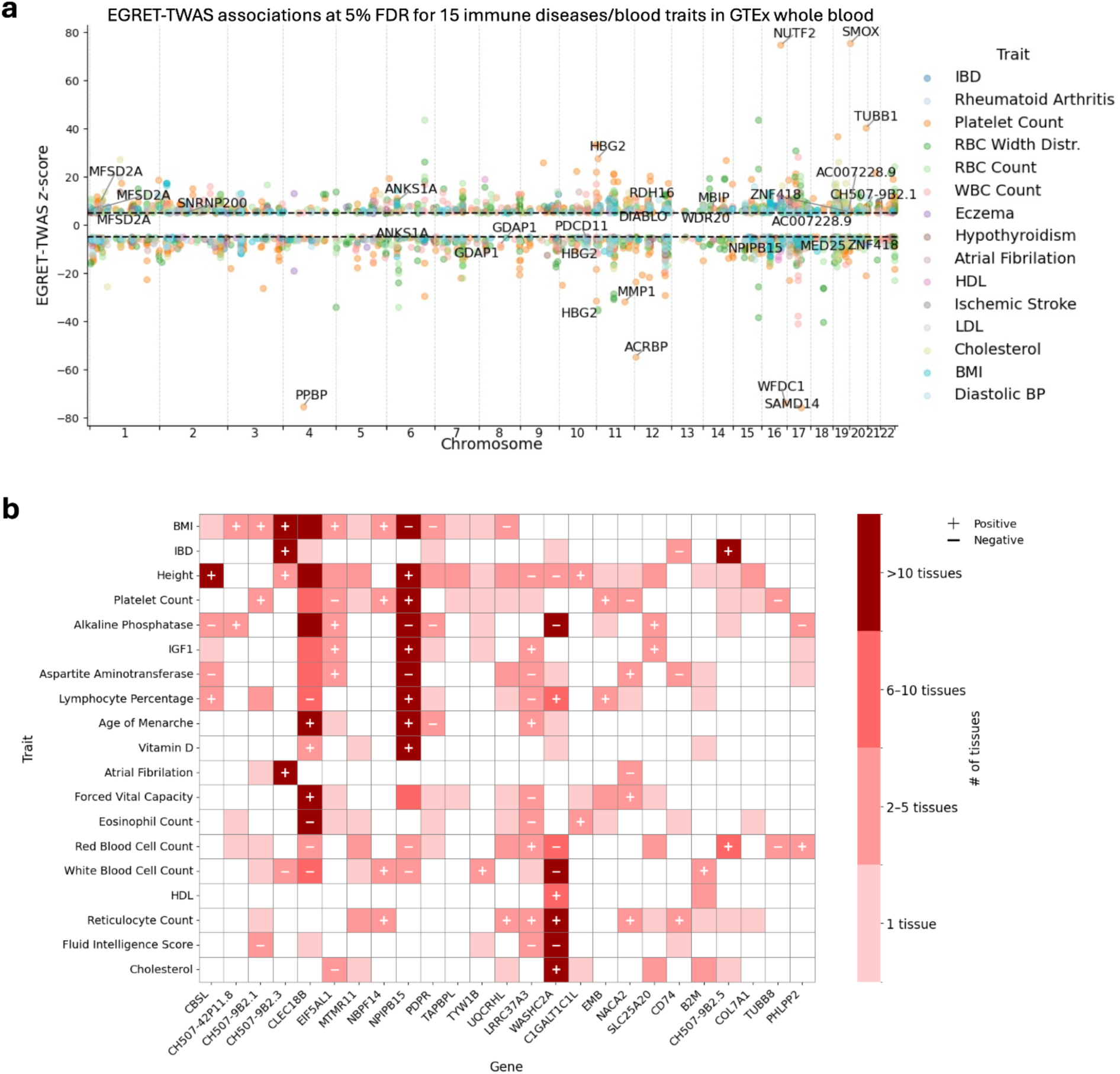
EGRET-TWAS framework colocalizes genome-wide effects on gene expression with disease-associated genetic variation. (a) Miami plot showing directional TWAS *z*-scores for EGRET whole blood gene models across a subset of 15 immune diseases and blood cell traits. Labeled associations were not identified by FUSION models and are *trans*-heritable. (b) We display the subset of the labeled genes from panel (a) that were associated with at least three complex traits/diseases. Colors are used to indicate the number of tissues harboring the same gene-disease association. Positive or negative signs indicate if the direction of gene-disease effect is consistently positive or negative across all associated tissues. Abbreviations: BMI, body mass index; IBD, inflammatory bowel disease; IGF1, insulin-like growth factor 1 hormone; HDL, high density lipoprotein. All numerical results can be found in **Supplementary Tables 7-8**.

Second, we discuss associations that are only detected with EGRET but not *cis*-FUSION models and are biologically plausible. *ANKS1A* is an example of a gene that is associated at 5% FDR with body mass index (BMI) and red blood cell (RBC) width distribution only with whole blood EGRET models but not FUSION models (EGRET nominal p = 9.71e-22, FUSION nominal p = 0.011). Recent work suggests that *ANKS1A* regulates LDL receptor-related protein 1 (*LRP1*), a cell surface receptor protein involved in lipid metabolism^67^. Further support comes from a study that found this gene to be associated with BMI using an independent whole blood eQTL dataset (the Young Finns Study^68^). Terrand and colleagues^69^ have extensively characterized *LRP1*’s involvement in lipid regulation. For example, *LRP1* is thought to act via the *WNT5A* signaling pathway to inhibit cholesterol accumulation. Additionally, variants within *ANKS1A* are linked to cardiovascular events in patients with familial hypercholesterolemia^70^. While the connection between *ANKS1A* and metabolic conditions is well established, its moderating effects on RBC width distribution is nascent. However, given its role in cholesterol regulation and being an integral component in cell membranes, it is plausible. Notably, RBC width distribution is linked to erythrocyte cell membrane cholesterol concentration and is considered a prognostic marker for cardiovascular disease^71^. Finally, we discuss associations that may indicate new biology not captured by previous studies. For instance, another finding implicates pro-platelet basic protein (*PPBP*) in platelet counts, which encodes the chemokine CXCL7 and has been linked to mitogenesis, extracellular matrix organization, and plasminogen activator synthesis^72^. We also identified *ZNF418* as significantly associated with IBD. *ZNF418* is a transcriptional repressor involved in the MAPK signaling pathway^721^ and may protect against gastric carcinoma^74^, a condition for which patients with inflammatory bowel disease (IBD) may be at increased risk^75^.

Lastly, we assessed the concordance of disease-gene associations across GTEx tissues. We restricted our analysis to 23,398 gene models that had a *trans*-genetic component and were newly associated via EGRET gene models (i.e., not via *cis* FUSION models) across each of 49 tissues and 78 traits. **Figure 5B** displays genes with at least one disease-specific association observed across at least 3 tissues and displays diseases/traits with at least one gene-specific association observed across at least 10 tissues (**Supplementary Table 8**). Several of these results are supported by previous studies. One such example is the association between cell surface receptor protein *CD74* and inflammatory bowel disease (IBD). While this gene has not been previously implicated by TWAS, other studies have linked it to intestinal inflammatory response^76,77^. As a second example, we investigate *WASHC2A* which is associated with many metabolic traits (HDL, cholesterol, alkaline phosphatase) and blood cell traits (white blood cell count, red blood cell count, reticulocyte count, lymphocyte percentage), as well as with height and fluid intelligence score, measuring cognitive functions across a range of tasks^78^. *WASHC2A* encodes a protein in the WASH complex involved in endosomal trafficking and actin regulation^79^. While few studies have directly studied the WASH complex in humans, ablation of *WASHC1* in mice resulted in elevated levels of plasma cholesterol^80^ with further studies showing that *WASHC1* plays a role in cholesterol absorption^81^, supporting the relevance of *WASHC2A* to cholesterol levels in humans. Concurrently, the WASH complex disrupts differentiation of hematopoietic stem cells and as a consequence influences white and red blood cell counts in murine model^82^ reinforcing its association across blood cell traits. Finally, as a more speculative case, we investigate *NPIPB15* which is associated with several blood traits (white blood cell count, red blood cell count, platelet count, lymphocyte percentage), several metabolic traits and markers (BMI, aspartate aminotransferase, alkaline phosphatase, IGF1, vitamin D), height and age of menarche. While *NPIPB15* encodes a nuclear pore-interacting protein (NPIP), its exact function is less well characterized but appears to be under high positive selection in the human population^83^. While there are no studies documenting its link to metabolic conditions, we investigated the upstream *trans*-regulators of *NPIPB15* as possible sources for these associations. Interestingly, we find that its *trans*-regulators, *IGFB2*, *TNIP3*, *ADGRG6*, *PHF14*, and *TRIM32,* have been previously associated with obesity-related conditions^84–88^. Altogether, we find that highly replicated gene associations using EGRET-TWAS nominate both biologically relevant disease biomarkers and plausible disease-critical genes which may be crucial to understanding underlying susceptibility to common complex traits and diseases.

### Gene regulatory networks characterized by shared long-range regulatory effects underlie disease susceptibility

Previous studies have demonstrated notable colocalization of GWAS loci and *trans*-eQTLs^18,20,^ suggesting that EGRET may help characterize networks of genes conferring disease susceptibility via eQTLs. We then hypothesized that genes sharing a common upstream *trans*-regulator may play cooperative roles in a particular biological process, which could be relevant for various polygenic diseases. To this end, we considered each tissue separately and leveraged the inferred *trans*-regulator genes of EGRET models, as discussed above for **Figure 4**, to construct a binary adjacency matrix. This matrix indicates the presence or absence of an EGRET weighted variant near potential upstream *trans*-regulators in the models of downstream target genes. We multiply the transpose of the adjacency matrix by itself to obtain a metric reflecting the coregulation of downstream genes. This matrix is then normalized and subtracted from 1 to obtain a distance metric between genes (**Methods**). Finally, we applied hierarchical clustering and applied a distance threshold of 0.9 to create gene modules. We only considered modules with at least 4 genes in the discussion below.

Across 49 GTEx tissues, we identified 78 total modules with an average of 5.65 genes per module (**Supplementary Table 9**). For 13 tissues, we did not identify any modules, likely due to lower sample size resulting in limited power. For thyroid tissue, we identified 8 modules of 5.25 genes on average, the largest value for a single tissue. Next, we set out to identify disease-critical modules. We hypothesized that if target genes were indeed regulated by an upstream regulator in a disease-critical manner, that EGRET-TWAS would produce stronger gene-disease associations on average than FUSION-TWAS using *cis*-eQTLs. To this end, we compared TWAS results from FUSION models and EGRET models for each of the 78 diseases/traits analyzed above.

We found that 1,407 unique module-disease pairs resulted in significantly greater EGRET-TWAS *z*-scores (one-sided paired t-test at FDR < 0.05) and that these results were largely tissue-specific. We discuss two notable examples here and summarize the other modules in the **Supplementary Table 10**. We refer to each module by its most frequently shared *trans*-regulator gene and use established regulatory mechanisms to hypothesize about how the genes work together to confer disease susceptibility or confer variation to complex traits. First, we discuss the *ARHGEF3* module in whole blood tissue consisting of 5 inhibited target genes and 5 activated target genes, of which most were strongly associated with platelet count (**Figure 6A**, **Supplementary Table 11**). This module exhibited the second strongest differential trait association between EGRET and FUSION of any module (**Figure 6B**, **Supplementary Table 11**). Averaged across 11 downstream target genes, EGRET-TWAS *z*-scores were significantly larger in magnitude relative to FUSION-TWAS (mean absolute EGRET-TWAS *z* = 45.56, mean absolute FUSION-TWAS *z* = 0.31, one-sided paired t-test p < 5.34e-04). The FUSION-TWAS *z*-score for all but 2 genes was zero, because for these genes FUSION could not construct a *cis*-eQTL gene expression prediction model with R^2^ > 0. EGRET finds significant associations between genome-wide eQTL models for 9 of these genes and platelet count at 5% FDR. *ARHGEF3* is a guanine nucleotide exchange factor, which is critical to many basic cellular processes and has long been known to activate the Rho/ROCK cascade and more recently has been observed to regulate platelet cell count^89,90^ (**Figure 6C**). This is consistent with our findings and may suggest a role for how *ARHGEF3* impacts downstream genes in a concerted effort to regulate platelet count. Intriguingly, we found nine of this network’s downstream genes to be modulated by a second upstream regulator *C3orf14* exclusively in whole blood tissue (**Figure 4C**); *C3orf14* is a transcription factor whose suppression is associated with acute myeloid leukemia^53^, a condition caused by the abnormal proliferation of hematopoietic stem cells and marked by low platelet coun^91^. Additionally, at least four of the genes in the module (*MMP1*, *TUBB1*, *PPBP*, and *LY6G6F*) have been independently linked to platelet-related traits^92–95^. Interestingly, several of the most strongly associated genes, e.g., *ACRBP*, *SMOX*, and *NUTF2*, have no known connection to platelet count but do play established roles in the immune system. *ACRBP* expression is associated with cytotoxic T cell^96^, *SMOX* is a chemokine suppressor^97^, and *NUTF2* suppresses immune cell infiltration and T cell receptor signaling in cancer^98^. Therefore, our analysis may suggest new roles for these genes in complex trait biology while also revealing that these genes may cooperatively work together.

**Figure 6.**
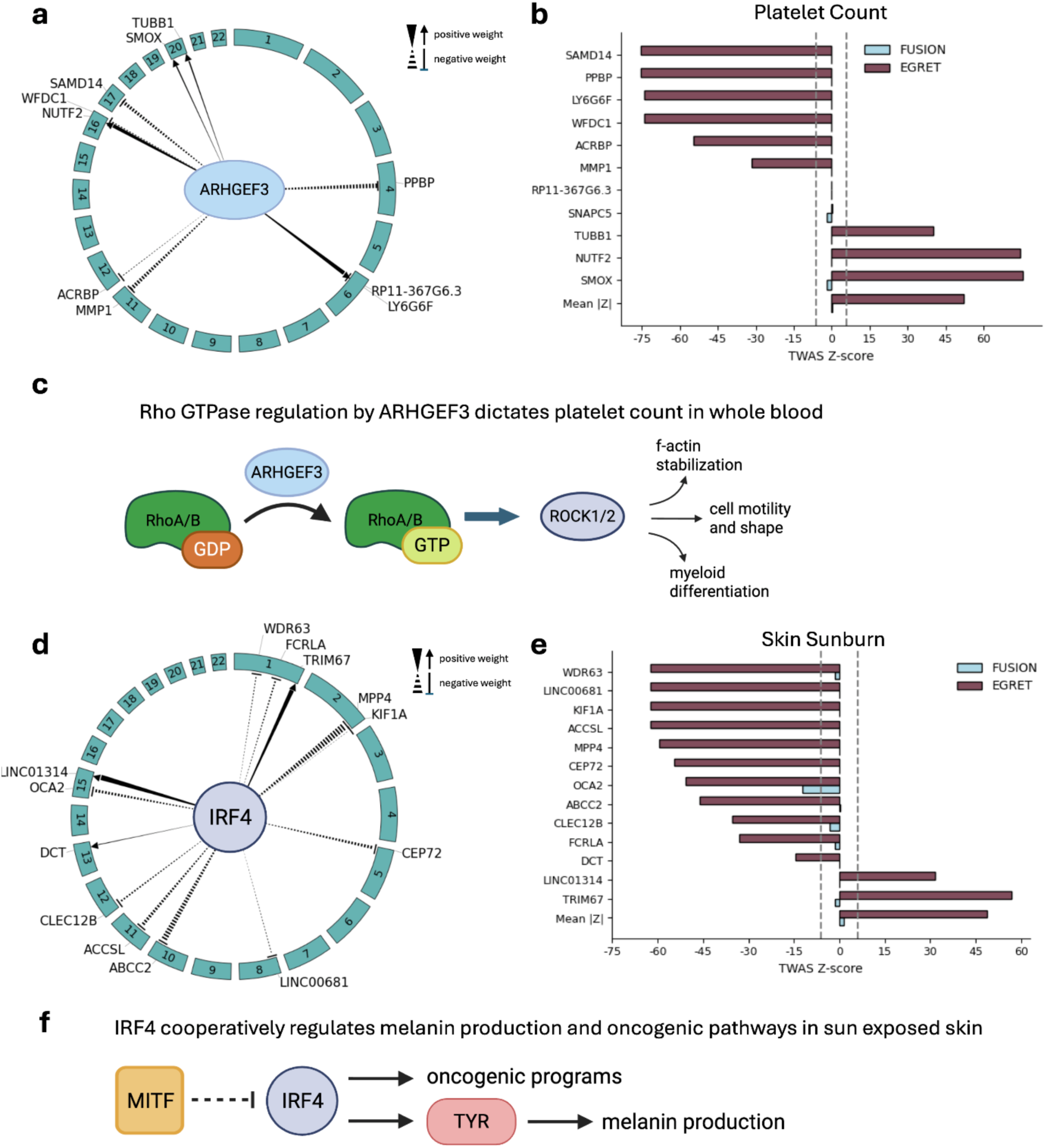
EGRET models key regulatory networks underlying complex traits and diseases. (a) *ARHGEF3* regulates downstream targets across the genome. Arrows indicate upregulation (positively weighted *trans*-variants) while blunt arrows indicate downregulation (negatively weighted *trans*-variants). (b) FUSION and EGRET TWAS *z*-scores for platelet count with genes in *ARHGEF3* module. (c) Schematic of established RhoA/B cascade regulation by ARHGEF3. (d) *IRF4* influences downstream genes across the genome. (e) FUSION and EGRET TWAS *z*-scores for sunburn with genes in the IRF4 module. (f) Schematic of established *IRF4* regulation potentially influencing genes involved in sunburn response. All numerical results can be found in **Supplementary Table 11**.

Second, we discuss the *IRF4* module in sun-exposed skin tissue consisting of 10 downregulated target genes and 3 upregulated target genes that are strongly associated with sunburn (**Figure 6D, Supplementary Table 11**). On average, the genes within this module were significantly more associated with sunburn using EGRET-TWAS (mean absolute EGRET-TWAS *z*-score = 48.6, mean absolute FUSION-TWAS *z*-score = 1.54, one-sided paired t-test p < 8.50e-08) (**Figure 6E**, **Supplementary Table 11**). FUSION-TWAS was unable to model the expression of 6 of these genes as they had no predictive model using *cis*-variants alone. *IRF4* (interferon regulatory factor 4) is a transcription factor with well-defined roles in skin pigmentation and acts as an oncogene in melanoma and myeloma, cancers of skin pigment cells (melanocytes) and aberrant B cell differentiation, respectively^99–101^. A large GWAS of skin tanning response and sunburn identified a variant located within an intronic enhancer of *IRF4* as a potential driver of these traits (p < 4.40e-157)^102^. Several of the downstream genes within the module are associated with melanin synthesis, the primary pathway responsible for skin color, including *OCA2* and *DCT* and are demonstrated to be regulated by *MITF*^103,104^ (**Figure 6F)**. While *MITF* is a known upstream repressor of *IRF4*^105^, *IRF4* has not independently been shown to have modulating effects on *OCA2* and *DCT,* potentially revealing a new role of this gene. Additionally, we found that there were many oncogenes within this module: *CEP72*, *TRIM67*, *KIF1A*, *ABCC2*, *CLEC2B*, *MPP4*, *LINC00681, and LINC0131*^106–113^. These highlight the alternative role *IRF4* may play as a major oncogene in various forms of melanoma, which is caused by the uncontrolled replication of melanocytes. One gene, *ACCSL,* has no prior connection to melanoma or skin pigmentation but is believed to function in amino acid metabolic processes as a paralog to *ACL*^114^, therefore, suggesting a new mechanism by which this gene may influence complex traits. We discuss other gene regulatory network examples in the **Supplementary Note**, such as one regulated by *GATA3* **(Supplementary Figure 14)**.

## Discussion

TWAS style approaches have been applied widely across gene expression cohorts and GWAS to nominate disease relevant genes^4,20,115^. However, most TWAS approaches model the effects of *cis*-eQTLs on genes^4,20^ even while studies have shown that gene expression regulation is largely orchestrated by *trans*-eQTLs^11–13^. Moreover, disease-critical effects are thought to act via *trans*-regulatory mechanisms^12,14,15^. As a result, traditional TWAS frameworks may be underpowered to detect disease relevant genes, as we showed in our simulation analyses. In this study, we developed EGRET, which extends standard *cis*-genetic gene expression models to include putative *trans*-eQTLs identified by state-of-the-art high-dimensional feature selection methods such as GBAT, *trans*-PCO, and Matrix eQTL. By training gene expression prediction models across 49 GTEx tissues, we show that through augmenting gene expression models with putative *trans*-regulatory variants, we can generate thousands more predictive gene models per tissue (R^2^ > 0, p < 0.01) compared to *cis*-eQTL models. Moreover, EGRET enabled the calculation of *trans*-genetic correlations of gene expression, and these appeared to be more tissue-specific than *cis*-genetic correlations. We also found that EGRET produced gene models with similar explained variance (R^2^) compared to state-of-the-art *trans*-eQTL gene expression prediction models generated by the BGW-TWAS and MOSTWAS frameworks, suggesting that while functional and statistical approaches differently select potential *trans*-regulatory variants, the increase in explained variance remains similar. Our study also highlighted 12,317 gene-tissue pairs with significantly nonzero *trans*-genetic components, which allowed us to dissect the individual contribution of GBAT, *trans*-PCO, and Matrix eQTL to EGRET models. These *trans*-heritable genes also resulted in many TWAS associations with complex traits/polygenic diseases that could not have been modeled by *cis*-eQTL frameworks. We also assessed the reproducibility of the *trans*-genetic regulation of gene expression by analyzing non-cross-mappable genes in benchmarking analyses and by validating that EGRET models capture *trans*-genetic variation in independent eQTL cohorts. Overall, the consistent proportions of explained variance (R^2^) by different *trans*-eQTL gene expression models and concordance of gene-disease associations in competing *trans*-TWAS frameworks reassures us that EGRET is capturing meaningful gene regulatory architecture. Lastly, we constructed gene regulatory networks based on shared *trans*-genetic components across genes and used these networks to understand how genes may collaborate to orchestrate disease susceptibility, supported by the concordance of gene-disease TWAS associations. These models additionally nominate disease-relevant genes that were previously not detected using models of *cis*-variants.

We note there are several limitations of this study, although we expect that our conclusions are robust to these limitations. First, as with any study of RNA-sequencing data, read mapping errors and multi-mapping due to sequence similarity could skew the normalized gene expression levels we use in our models^19^. Second, although we aim to reduce overfitting by regularization, cross-validation, and evaluating the predictive power of EGRET models in an independent eQTL cohort, the current dearth of population-level RNA-sequencing datasets in non-blood tissues makes it challenging to verify that EGRET is capturing reproducible *trans*-regulatory loci out-of-cohort. Third, while *trans*-eQTLs and gene regulatory networks are highly cell-type specific, constructing EGRET models in bulk tissues may result in diluted detection of *trans*-genetic components. Rather, applying EGRET to cell-type-specific bulk RNA-sequencing or single cell RNA-sequencing may result in strong *trans*-genetic correlations across cell types due to noise reduction, more variance explained of gene expression levels, and more gene-disease associations via TWAS. Lastly, in bulk tissue RNA-sequencing, cell type proportions (or composition) can vary across individuals and subsequently be associated with genetic variation across the entire genome. This phenomenon predominantly causes cell-type-specifically expressed genes to appear associated with genetic variation. Without knowing the cell-type-proportions for these individuals or having a reliable deconvolution approach to infer those proportions, resolving this limitation is challenging. Despite these limitations, EGRET presents a powerful strategy for modeling genome-wide genetic variants and discovering novel gene-trait associations and regulatory programs underlying disease.

## Methods

### Components of genome-wide EGRET model

First, EGRET models all SNPs within a 1 megabase (Mb) window centered at the transcription start site (TSS) as *cis*-predictors of gene expression by default. This baseline model is mathematically equivalent to FUSION^4^, where the expression level of a single gene across individuals is modeled as a weighted linear combination of allele dosages:

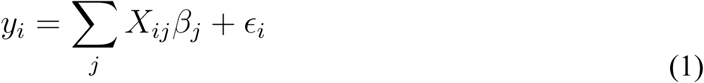

where for a given gene, *y_i_* is the normalized gene expression for individual *i*, *X_i,j_* is the allele dosage for individual *i* at variant *j*, *β*_j_ is the effect size of *cis*-variant *j* on expression, *ɛ_i_* is the residual noise not explained by *cis*-genetic effects, which is assumed to be normally distributed.

Second, EGRET applies high dimensional feature selection strategies to all SNPs beyond the 1 Mb *cis* window to identify putative *trans*-eQTLs, namely Matrix eQTL^24^, GBAT^19^, and *trans*-PCO^17^, each described in detail below.

Matrix eQTL (v. 2.3) is a computationally efficient framework for conducting association tests for billions of SNP-gene pairs. We tested approximately 8.5 billion SNP-gene pairs per GTEx tissue. Matrix eQTL tests for a linear relationship between the dosage of a SNP and normalized gene expression level across individuals using linear regression. For all association tests using GTEx data, we regress out standard covariates provided by the GTEx consortium including the top 5 genotyping principal components, tissue-specific gene expression PEER factors ranging from 15-60 factors, age, sex, and sequencing platform. We additionally use Matrix eQTL to enable GBAT and *trans*-PCO analyses, discussed below. In all applications, we use the general linear model option as specified by the parameter useModel = modelLinear. For each *trans*-eQTL association strategy, we only retain variants with FDR < 10% in at least one strategy.

EGRET utilizes a modified approach to the Gene Based Association Test (GBAT) introduced by Liu and colleagues^19^. The goal of GBAT is to find *cis*-eQTLs mediating *trans*-regulatory relationships between genes. GBAT first trains *cis*-genetic models of expression for each gene using cvBLUP^116^ and then attempts to associate the imputed expression for a focal (or upstream) gene with measured expression levels of downstream genes. Specifically, in order to make *cis*-genetic predictions across the GTEx cohort, we used *cis-*genetic gene expression models trained using FUSION to impute *cis*-genetic prediction values across individuals instead of cvBLUP; we elected to make this change due to the computational costliness of running cvBLUP across 49 GTEx tissues, because LASSO would prioritize fewer variants into our feature space, reducing the risk of overfitting and over-parametrizing our model, and because LASSO has similar predictive performance as BLUP models (**Supplementary Figure 6**). Focusing only on genes with a significantly positive R^2^ (p < 0.01) from the CV LASSO model, we associated each gene’s predicted expression with the normalized expression level of each gene that is at least 5 Mb away from the focal gene. Our second modification of the method was to perform gene-gene association testing using Matrix eQTL due to its computational efficiency, rather than a standard association test in R. We applied multiple hypothesis correction using FDR across the number of upstream *trans*-regulator genes considered. Of FDR < 10% significant gene-gene pairs, the variants with non-zero weight in the LASSO model for the upstream focal gene are considered as putative *trans*-eQTLs (or features) in the EGRET model for the downstream gene. We note that the original GBAT tool was not designed for variant-level *trans*-eQTL discovery, but rather broader *trans*-regulatory loci mapping.

The final component of EGRET is a modified implementation of *trans*-PCO^17^. *trans*-PCO was developed for detecting *trans-*effects on gene regulatory networks. First, *trans*-PCO uses WGCNA clustering to define co-expressed gene modules. Next, it tests the linear association between a single genetic variant and the module’s expression using a principal component omnibus (PCO) test, which combines six PC-based association tests to boost power^17^. Co-expression clustering is done using WGCNA and only 10 PEER factors, as in the original *trans*-PCO study, for each tissue as potentially higher order PEER factors may capture large-scale *trans*-regulatio^37^. Next, we use Matrix eQTL to generate summary statistics across all modules and genome-wide SNPs. Finally, we follow the same 6 PC-based multivariate association test procedure used previously to determine the final multivariate p-value association statistic. We retain only the variant-module pairs with FDR < 10%, correcting for the number of genome-wide variants tested. In downstream EGRET model fitting, significantly associated variants are included as putative *trans*-eQTLs in the models of all genes represented by the module, excluding genes that are on the same chromosome as the variant.

### EGRET Model Training

EGRET takes the union of prioritized variants from the *cis* window, Matrix eQTL, GBAT, and *trans*-PCO, as discussed above:

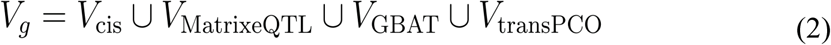

EGRET fits 4 different linear models using the union of *cis* and *trans* prioritized variants, *V_g_*. A variant does not receive any higher priority if it is identified by more than one of these methods. Thus, the complete genome-wide EGRET model is represented by the following linear combination of *cis* and *trans* variants.

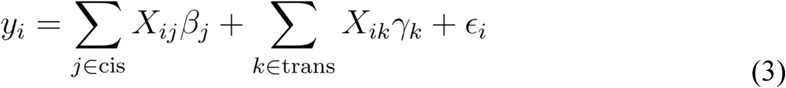

Here, equation (3) builds on equation (2) by letting *k* index *trans*-variants. *X_i,j_* is the allele dosage for individual *i* at variant *k*, *γ*_k_ corresponds to the effect sizes of *k* putative *trans*-eQTLs in the model, and *ɛ_i_* is the residual noise not explained by genome-wide genetic effects which is normally distributed.

Three of the model fitting strategies (LASSO, elastic net, and BLUP) are shared with FUSION. In order to leverage information regarding the source of the putative *trans*-eQTLs, we add an additional baseline model, xtune LASSO^31^, that learns priors based on the membership of variants to different model categories using an empirical Bayes approach and adjusts the lambda penalty term in accordance with this. This prior information is encoded by a matrix with *n* rows, e.g., the number of variants in the model, and *m* columns, e.g., the 3 *trans* mapping methods.

Variants receive a value of 1 for the methods that found an association between genotype and gene expression at FDR < 10%, otherwise a zero is recorded. All methods are trained in a 5-fold cross validation scheme, where the regressed expression of samples from the training fold are fitted against the standardized genotypes of those samples.

One goal of EGRET is to identify which genes are significantly regulated by *trans* loci. Splitting the genome-wide models further into *cis*-variant and *trans*-variant models helps us interpret the genetic architecture of a gene’s expression. Given a coefficient or weight vector, *w*, the *cis*-only variant model is constructed from weighted variants which lie within an extended *cis* region (+/-2.5 Mb of the TSS); this means that the weights of variants that lie between the original 1 Mb *cis* region and the extended 5 Mb *cis* region are generated from one of the *trans*-eQTL association methods. The *trans*-only variant model is defined exclusively by variants that have non-zero weights and are either off chromosome or outside of the extended *cis* region on the same chromosome. Genes whose *trans*-only variant models have a significantly positive R^2^ in CV (p < 0.01) are referred to as *trans*-heritable in the main text. Finally, the genome-wide model uses all weighted variants during model fitting. This workflow helps to minimize computational resources as only one genome-wide model is fit and separate genomic region models are evaluated for their performance.

### Simulations

We designed a simulation framework to evaluate EGRET across two major dimensions: (1) its ability to capture *cis*- and *trans*-eQTL effects in gene expression prediction, and (2) its performance in nominating disease-relevant genes through TWAS compared to *cis*-eQTL only models like FUSION.

To address the first goal, we used real genotype data from 715 European individuals in GTEx and simulated gene expression as a function of both *cis*- and *trans*-eQTLs, plus environmental noise as proposed in Wang et al. 2023^117^. For the second, we simulated both genotype and complex trait data, generating expression-based traits and assessing whether EGRET-TWAS improved power for gene discovery compared to FUSION, while maintaining a well-calibrated false positive rate.

To create a feasible and realistic simulation framework, we first selected appropriate genomic regions and genes. To improve computational efficiency when constructing LD matrices, we identified genes with at least 200 variants but no more than 2,000 variants in a 1 Mb *cis* window. From these genes, we randomly selected 100 genes per simulation experiment.

We simulated gene expression using real genotype data from GTEx individuals and chosen causal effect sizes. Expression for individual *i* was modeled as a linear combination of known causal *cis*-eQTL and *trans*-eQTL effects (**β*_j_,γ*_k_, respectively). Causal effect sizes *β* were sampled from a normal distribution *N(0,σ^2^_ꞵ_)*, with the variance determined by the per-SNP heritability: *σ^2^_ꞵ_ = h^2^/k*, where *h^2^* is the total genome-wide heritability and *k* is the number of causal SNPs (default: 5 *cis*-eQTLs, 20 *trans*-eQTLs per gene). While the default number of *cis*-eQTLs was selected based on empirical eQTL fine-mapping analysis of the GTEx cohort^25^ and fine-mapping analyse^118^, we varied the number of *trans*-eQTLs between 5 and 50 to evaluate EGRET’s robustness across different *trans*-regulatory architectures. *cis*-eQTLs were sampled from within +/- 500 kb of the TSS of the gene of interest, while *trans*-eQTLs were selected randomly from the *cis* windows of other genes on other chromosomes to avoid LD contamination.

For simulated data, we exclusively used Matrix eQTL to train EGRET gene expression models, as this method does not require specific regulatory structure across other genes and is suitable for simulated data without gene-gene co-regulation. For benchmarking, we built FUSION models using only *cis*-variants (within +/- 500 Kb of the TSS). For both methods, the maximum CV R^2^ from the multivariate regression frameworks was recorded for each gene, and we computed the mean and standard error of this metric across all simulated genes.

To assess the utility of EGRET and FUSION models in TWAS, we simulated a complex trait influenced by gene expression. To this end, we simulated a 50,000 person GWAS cohort by generating genotypes based on the LD structure of the GTEx cohort. These genotype positions were restricted to the *cis* regions of the genes above. We treated all other genotype positions beyond these *cis* windows as noise and thus modeled their GWAS *z*-scores with a normal distribution with mean 0 and variance 1. The complex trait *Y_i_* for each individual was modeled as a function of the genetic component of gene expression, *y*_i_, from genome-wide eQTLs:

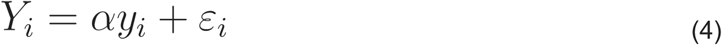

where *α* was sampled from a *N(0,1)* distribution and noise was added such that 10% of complex trait variance is explained by the genetic component of expression. 10% of genes were selected to be causal at random. GWAS summary statistics were then computed using linear regression, adjusting for the top 5 genotype principal components.

TWAS *z*-scores were computed using a weighted burden test, as described in the next section, where SNP-level GWAS *z*-scores were integrated with weights from each expression model. We assessed performance in terms of power and false positive rate using a nominal threshold of p < 0.05 and generated AUROC curves across 15 p-value cutoffs between 0 and 1.

### EGRET-TWAS with Summary Statistics

We utilize the general framework presented in FUSION-TWAS summary statistics given by the formula below. Here, Z is a vector of *z*-scores of SNPs from the simulated GWAS with dimension m representing each of the variants that have a non-zero weight in the genome-wide EGRET model. W is the corresponding weight vector also with dimension m from the EGRET model. Σ_s,s_ is the LD or covariance matrix of the m SNPs generated from genotype data from 489 European individuals from the 1000 Genomes Project as the LD reference panel.

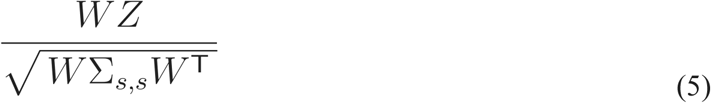

### Competing methods for gene expression prediction

We compare the performance of EGRET against the *cis*-eQTL FUSION^4^ model and two alternative genome-wide methods, MOSTWAS^21^ and BGW-TWAS^22^. We note that we do not compare EGRET to Predixcan^20^, another widely used *cis*-eQTL method, as it has been shown to perform similarly to FUSION^119^.

FUSION models are trained using *cis*-variants within a +/- 500 kb region centered on the TSS of the gene. FUSION considers 5 multivariate linear approaches, e.g., LASSO, elastic net, BLUP, Top1, and BSLMM. The model with the largest CV R^2^ is selected. We benchmarked EGRET with FUSION across all protein coding, lincRNA, and anti-sense RNA genes expressed in each of the 49 GTEx tissues. We run TWAS on FUSION models as previously described^4^, which is similar to the summary statistics approach described in the previous section. Disease-associated genes are selected based on FDR < 0.05, correcting for the number of tested gene models (CV R^2^ p < 0.01).

We also benchmarked EGRET against MOSTWAS and BGW-TWAS, two methods that aim to include genome-wide variants into gene models. We run both with default parameters across 4,854 non-cross-mappable genes in the GTEx whole blood tissue.

### MOSTWAS

MOSTWAS^21^ is an ensemble approach similar to EGRET which trains genome-wide models of expression using two approaches, mediator-enriched TWAS (MeTWAS) and distal-eQTLs mediated by local biomarkers (DePMA). MeTWAS identifies mediating biomarkers such as microRNAs, CpG sites, and transcription factors that may influence gene expression from across the genome. For this method, we test the mediating effects of all non-cross-mappable genes which fall under protein coding, lincRNA, and antisense RNAs. In their second approach, DePMA attempts to find mediating effects of distal-eQTLs on biomarkers local to these SNPs.

We apply both methods in accordance with their default recommended settings. The maximum CV R^2^ between the MeTWAS and DePMA models was recorded for each gene.

### BGW-TWAS

Bayesian Genome-wide TWAS (BGW-TWAS) employs Bayesian variable selection regression and the scalable expectation-maximization Markov Chain Monte Carlo (EM-MCMC) algorithm to build genome-wide models of expression^22^. We applied their software with default parameter settings. Since BGW-TWAS does not natively supply a CV R^2^, we train the model on 80% of GTEx whole blood samples and use the R^2^ of the test set, e.g., the remaining 20% of samples, to assess the model’s performance.

### Determining trans-regulators for each tissue

To identify the potential upstream genes harboring *trans* loci responsible for improving the prediction of our *trans*-heritable genes, we used eQTL summary statistics from GTEx v8^7^ to identify if our weighted *trans*-variants are implicated as *cis*-eQTLs of any of their neighboring genes, which we later refer to as putative *trans*-regulators. For weighted *trans*-variants that were not listed as a *cis*-eQTL of a gene, we defaulted to use the nearest expressed gene in that tissue as the putative *trans*-regulator and reasoned that such a variant could impact functional mechanisms such as splicing or protein-protein interactions of that nearest gene.

### Enrichment analysis procedure

Enrichment of *trans*-regulators and *trans*-weighted variants in EGRET models was done using a hypergeometric test to evaluate over or underrepresentation in certain functional categories. For gene enrichment, we use gene ontology from UCSC table browser for the table called *gtexGeneV8* in conjunction with transcription factors described by Lambert and colleague^609^. We restrict the transcription factors to those with experimentally demonstrated DNA binding specificity. This excludes co-factors and RNA binding proteins. For variant enrichment we use annotation data from two different studies. For GTEx *cis*-eQTL enrichment, we use GTEx v8 summary statistics^25^ for the respective tissue. For other genomic annotations, we use v2 of the baseline LD from LD Score regression^6^ (**Data availability**).

### Construction of trans-co-regulation regulatory networks

One major goal of EGRET is to elucidate genetic regulatory networks underlying complex traits and diseases. To do this, we build gene networks using the *trans*-regulators we defined previously. First, we create a binary adjacency matrix, called *M*, representing the connection between target genes and the upstream genes regulating them. In this matrix, a 1 represents a regulatory relationship defined by the previous section. To determine genes that are co-regulated we compute M.T x M which results in G, a matrix where each entry is the number of shared regulators between genes and thus quantifies coregulation. The matrix is then normalized so that diagonal entries equal 1 given by the following formula:

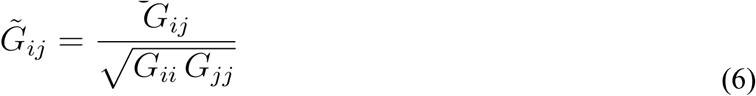

Finally, we compute 1 - G to form a distance matrix for clustering. We then cluster this matrix using the fcluster() function in *scipy* with a distance threshold of 0.9. We also evaluated clustering across other distances and found that the 0.9 threshold allows us to permissively assign genes to co-regulation clusters, which is desirable as our binary input matrix does not indicate the strength of co-regulation but rather the presence of co-regulation. Finally, we select clusters of genes with at least 4 genes and rank genes by how frequently they regulate the other genes in the cluster based on the *trans*-regulator connections we identified previously.

### Tissue co-expression analysis within GTEx

To assess shared regulatory effects across tissues in the *cis* and *trans* regions, we used the 1000 Genomes Project^120^ to impute expression from EGRET models trained within the 49 GTEx tissues^25^. First, since calculating genetic correlations is not straightforward when effect sizes are estimated with sparse regularization models, we compute correlations of imputed expression by taking the product of EGRET model weights *β_EGRET_* (separated into *cis-* and *trans-*SNPs (> 5 Mb) from the gene TSS) and the standardized genotypes of 489 European-ancestry individuals from the 1000 Genomes Project, *X_1KG_*, where the rows index individuals and the columns index SNPs. This can be formalized by the equation below.

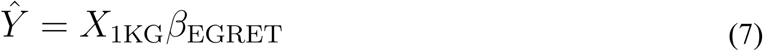

For a given pair of tissues, we report the average of these Pearson correlations across the set of genes with a significantly predictive model (R^2^ > 0, p < 0.01) in at least one of the tissues. If a model doesn’t exist or is not significantly predictive in one of the tissues, the Pearson correlation for that gene is set to be 0 across those two tissues.

### Replication in DGN dataset

To perform robust replication of GTEx-trained EGRET models in DGN, we imputed the genotypes of DGN individuals using the Michigan imputation server with 1000 Genomes phase 3 v5 reference panel and Eagle v2.4 phasing engine. Then, we selected *trans*-heritable genes (R^2^*_trans_* p < 0.01) to assess the utility of GTEx models applied out of cohort. Finally, to assess the contribution of *trans*-variants modeled by EGRET we compared the predictive accuracy of exclusively *cis*-variants and the genome-wide model which includes *trans*-variants.

### DGN dataset processing

We preprocessed DGN RNA-sequencing data similarly to GTEx RNA-sequencing data. We converted read counts to transcripts per million (TPM) and then quantile normalized gene expression levels across individuals. Finally, we apply an inverse normal transformation across individuals as done previously^25^.

### Processing individual-level genotypes

All genotype preprocessing is done using PLINK v2.0. Variant filtering includes removing variants that have a minor allele frequency (MAF) < 0.05. For pairs of variants in high LD within a 100 kb window, we used the PLINK --indep-pairwise flag to retain only one representative variant in order to retain a high density of variant markers but simultaneously remove highly redundant features^121^.

### Cross-mappable region pruning

Cross-mappable regions are a well-established confounder in *trans*-eQTL analyses and can create false-positive associations in which apparent *trans*-associations may reflect mapping artifacts rather than true regulatory effects^23^. To mitigate this problem, we systematically removed variants located in regions (of gene A, referred to below) with high cross-mappability relative to the target gene (referred to as gene B below). We leverage precomputed gene-level cross-mappability scores from Saha and Battle^23^. In their framework, cross-mappability from gene A to gene B, denoted crossmap(A, B), is defined as the number of k-mers from gene A that align within gene B. By default, k-mer lengths are 75 bp for exonic regions and 36 bp for UTR regions. When reads from gene A misalign to gene B due to sequence similarity, gene B’s measured expression may partially reflect transcripts from gene A. This can generate spurious *trans*-eQTL signals in which variants near gene A appear associated with expression of gene B.

To account for this, for each gene B for which we construct genome-wide EGRET models, we identify genes whose sequences are highly cross-mappable to gene B. Specifically, we first compute the background cross-mappability rate for gene B by summing the total number of reads from all other genes that are predicted to misalign to B and then dividing by the number of genes considered. This yields the average number of misaligned reads per gene targeting B. We then classify a particular gene A as highly cross-mappable to gene B if crossmap(A, B) exceeds this background average. For each such gene A, we define a 100 kb window centered on its transcription start site (TSS). We then selectively exclude variants that fall within this window, before constructing the EGRET model for gene B.

### Statistics and Reproducibility

First, to ensure robustness of interpretations from our simulation analyses, we simulated 100 independent genes for each genetic architecture to evaluate EGRET performance. Second, as described above, we subjected genotyping data to several filters to ensure we are only analyzing common variants (minor allele frequency > 5%) and pruned variant pairs that are in high LD (R^2^ > 0.9^121^ within a 100 kb window, keeping only one representative variant, in order to retain a high density of variant markers but simultaneously remove highly redundant features. Third, we transformed expression data using quantile normalization and the inverse normal transformation. Fourth, we only analyzed genes that are protein coding, lincRNAs, or antisense to reduce the chance of spurious associations with genes without established functional roles, such as pseudogenes, microRNAs, small nucleolar RNAs, and intronic transcripts. Fifth, we only conducted TWAS on genes with a CV R^2^ p < 0.01 to prevent spurious complex trait/disease associations with noisy gene expression prediction models. Randomization and blinding were not pertinent to our study.

### Data Availability

GTEx gene expression and genotype data were acquired from dbGaP accession phs000424.v9.p2. GEUVADIS genotype data is publicly available at https://www.ebi.ac.uk/biostudies/arrayexpress/studies/E-GEUV-1. 1000 Genomes LD reference files were acquired from https://www.bridgeprs.net/guide_input/. DGN gene expression and genotype data was acquired from the NIMH (National Institute of Mental Health) Repository and Genomics Resource. Genomic annotation files from the baseline LD model can be found at https://alkesgroup.broadinstitute.org/LDSCORE/. EGRET gene models and EGRET-TWAS association statistics can be downloaded from Zenodo at https://zenodo.org/records/18718581.

### Code Availability

EGRET software including documentation and tutorial is publicly available at https://github.com/AmariutaLab/EGRET [10.5281/zenodo.18718581]. The Mancuso Lab TWAS Simulator is available at https://github.com/mancusolab/twas_sim. FUSION software is available at http://gusevlab.org/projects/fusion. Matrix eQTL software is available at https://github.com/andreyshabalin/MatrixEQTL. GBAT software is available at https://github.com/xuanyao/GBAT. trans-PCO software is available at https://github.com/liliw-w/Trans. MOSTWAS software is available at https://github.com/bhattacharya-a-bt/MOSTWAS. BGW-TWAS software is available at https://github.com/yanglab-emory/BGW-TWAS.

## Acknowledgements

This work was supported by funding from the National Science Foundation (NSF) (Award #2336469 awarded to T.A.) and the National Institutes of Health (NIH) (NHGRI R01HG013671 awarded to T.A.). The funders played no role in study design, data collection and analysis, decision to publish or preparation of the manuscript. We would like to thank the GTEx and DGN cohort participants. The Genotype-Tissue Expression (GTEx) Project was supported by the Common Fund of the Office of the Director of the National Institutes of Health, and by NCI, NHGRI, NHLBI, NIDA, NIMH, and NINDS. This work used the Expanse HPC server at the San Diego Supercomputer Center (SDSC) through allocation BIO230210 from the Advanced Cyberinfrastructure Coordination Ecosystem: Services & Support (ACCESS) program, which is supported by National Science Foundation grants #2138259, #2138286, #2138307, #2137603, and #2138296.

## Author contributions

K.B. and T.A. conceived and designed the study. K.B. conducted simulation analyses and real data analyses. M.F.R. helped design figure panels. K.B., M.F.R., and T.A. wrote the initial draft of the manuscript and contributed to the final manuscript.

