## Supplementary Note for "Leveraging genome-wide effects on gene expression to identify disease-critical genes with *trans*-genetic components"

In addition to the gene regulatory networks discussed in the Results subsection *Gene regulatory networks characterized by shared long-range regulatory effects underlie disease susceptibility,* here we discuss the *GATA3* module in cultured fibroblasts consisting of 10 downregulated, or inhibited, target genes and 1 upregulate, or activated, target gene which are modestly enriched for eczema and atopic dermatitis GWAS loci (**Supplementary Figure 14**). Notably, the gene body of *GATA3* harbors a highly significant eczema GWAS locus (lead SNP $p < 9.61e\text{-}08$)[122], and therefore we hypothesized that downstream targets of *GATA3* may also be associated with eczema. While only one downstream gene received a statistically significant EGRET-TWAS association, considering an average across all 11 downstream target genes, EGRET-TWAS *z*-scores were indeed significantly larger in magnitude relative to FUSION-TWAS (mean absolute EGRET-TWAS *z*-score = 2.33, mean absolute FUSION-TWAS *z*-score = 0.12, one-sided paired t-test $p < 0.0125$). Considering the genes under *GATA3* regulation, we find that they share common biological functions related to cell cycle regulation (GO:0051726 FDR < 2.82e-07), implicating specifically the genes *KIF14*, *ASPM*, *AURKA*, *CDC20*, *BUB1B*, *CENPA*, *SGO2*, *FAM83D*, and *CKS2.* Intriguingly, many of these genes harbor FOXM1 transcription factor binding sites. *GATA3* is known to regulate *STAT3*[123], which in turn is known to regulate *FOXM1*[124], which may then regulate the genes we identified by our module-based analysis. *FOXM1* plays a critical role in fibrogenesis and cell proliferation in cancers and fibrotic disorders[125,126]; but its specific role in eczema has not been reported before. However, fibrogenesis causes tissue remodeling in individuals with eczema[127,128], which in turn supports the association we find between *MSTN* and eczema. *MSTN* promotes the proliferation of fibroblasts and their further differentiation into myofibroblasts which drive fibrosis[129]. Overall, our findings highlight the utility of modeling genome-wide genetic effects on gene expression as a way to characterize the interplay between genes as they work cooperatively to confer disease susceptibility and regulate complex traits.

### Supplementary Figures

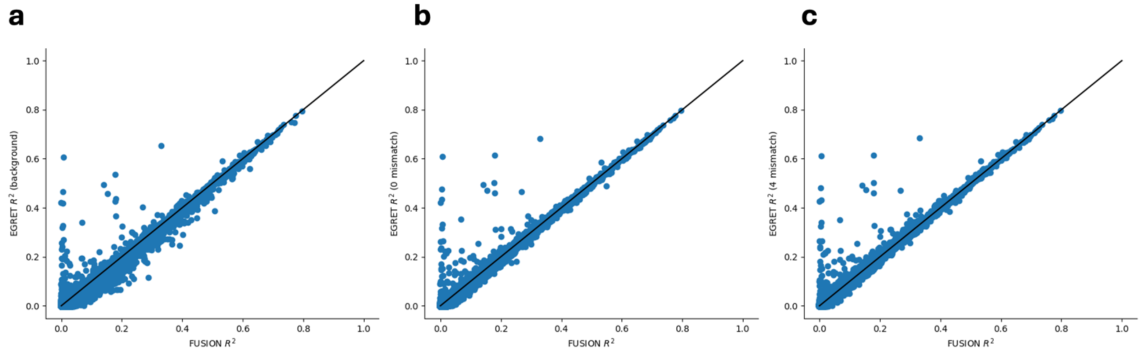


**Supplementary Figure 1. Cross-mappability thresholds.** Performance ($R^2$) of Matrix-eQTL only EGRET models (y-axis) compared to FUSION model performance (x-axis) across all genes with varying cross-mappability thresholds applied. (a) Background cross-mappability determined empirically. (b) Allowing for 0 misaligned reads between target gene and other genes. (c) Allowing up to 4 misaligned reads between the target gene and other genes. Black lines denote the y = x line. Overall we find that results are not sensitive to the threshold selected.

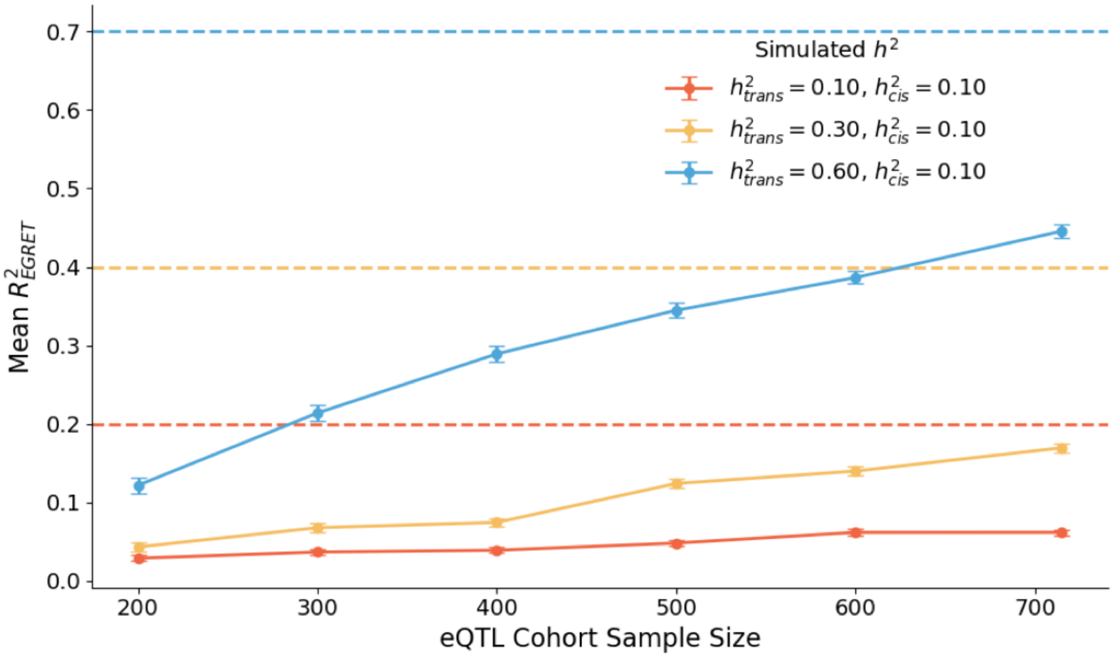


**Supplementary Figure 2. Comparison of gene model performance.** Performance of Matrix eQTL-only EGRET in capturing genome-wide heritability of gene expression at different eQTL cohort sizes (ranging from 200 to 715 individuals to represent relevant GTEx sample sizes) and at different combinations of *cis*- and *trans*-heritability. Error bars represent standard errors across 100 independent simulations at each parameter setting. By default, we simulate 5 *cis*-eQTLs and 20 *trans*-eQTLs. Increasing the sample size has a more profound effect on the model performance (CV $R^2$) at larger *trans*-heritabilities than smaller *trans*-heritabilities. Dashed lines indicate total genome-wide heritability, calculated additively between *cis*- and *trans*-eQTLs.

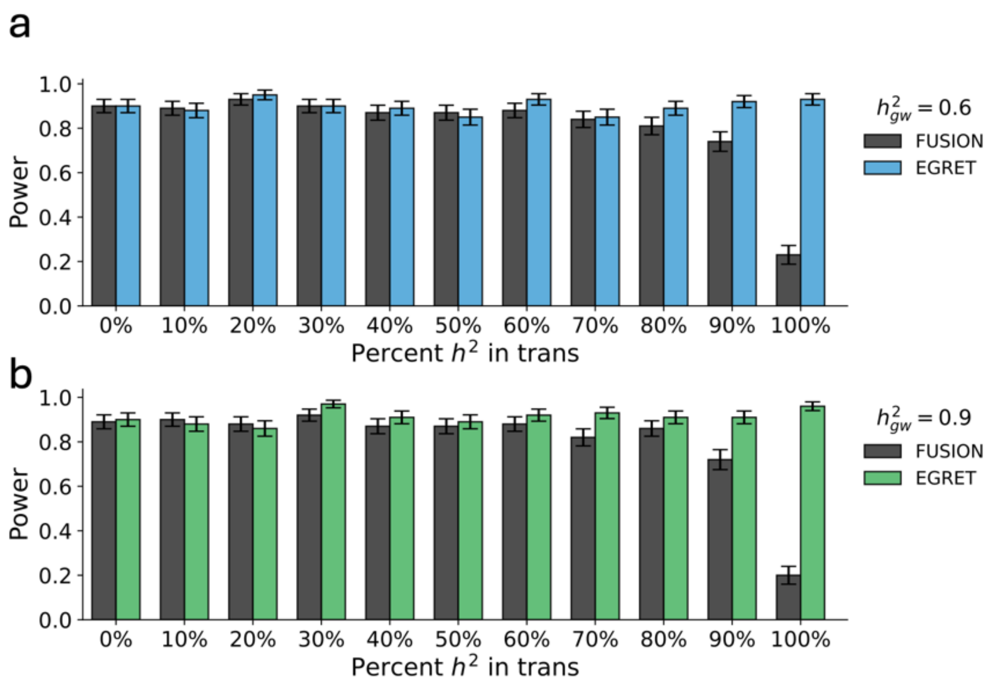


**Supplementary Figure 3. Comparison of TWAS power.** TWAS power to identify disease-critical genes at nominal significance using Matrix eQTL-only EGRET and FUSION models at different proportions of *cis*- and *trans*-heritability of gene expression. Error bars represent standard errors across 100 independent simulations of each parameter setting. When the genome-wide heritability = 60% (a) or genome-wide heritability = 90% (b), we exhibit similar differences as when genome-wide heritability = 30%, as shown in **Figure 2C**. Genetic effects are distributed by default in 5 *cis*-eQTLs and 20 *trans*-eQTLs. Gene models are trained in 715 individuals, 50K GWAS individuals are simulated with gene expression effects accounting for 10% of trait heritability.

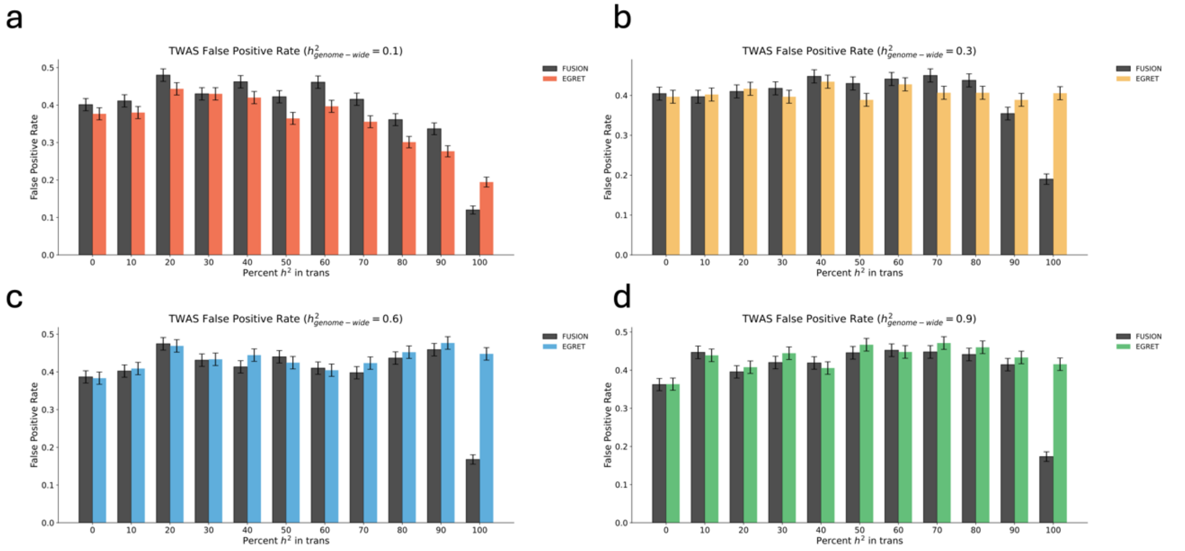


**Supplementary Figure 4. Comparison of TWAS false positive rates.** TWAS false positive rate using EGRET and FUSION models at different genome-wide heritabilities: (a) 10%, (b) 30%, (c) 60%, (d) 90%. Error bars represent 95% confidence intervals across 100 independent simulations of each parameter setting. Genetic effects are distributed by default in 5 *cis*-eQTLs and 20 *trans*-eQTLs. Gene models are trained in 715 individuals, 50K GWAS individuals are simulated with gene expression effects accounting for 10% of trait heritability.

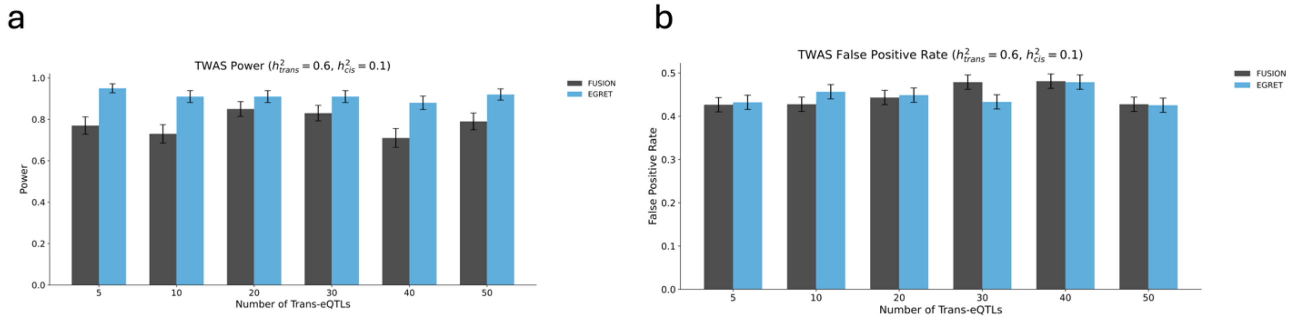


**Supplementary Figure 5. TWAS power and false positive rates.** TWAS power (a) and false positive rates (b) using EGRET and FUSION models across a range of simulated causal *trans*-eQTLs (from 5 to 50) where ~85% of genome-wide heritability resides in *trans* regions. Error bars represent 95% confidence intervals across 100 independent simulations of each parameter setting. Genetic effects are distributed by default in 5 *cis*-eQTLs and 20 *trans*-eQTLs are simulated. Gene models are trained in 715 individuals, 50K GWAS individuals are simulated with gene expression effects accounting for 10% of trait heritability.

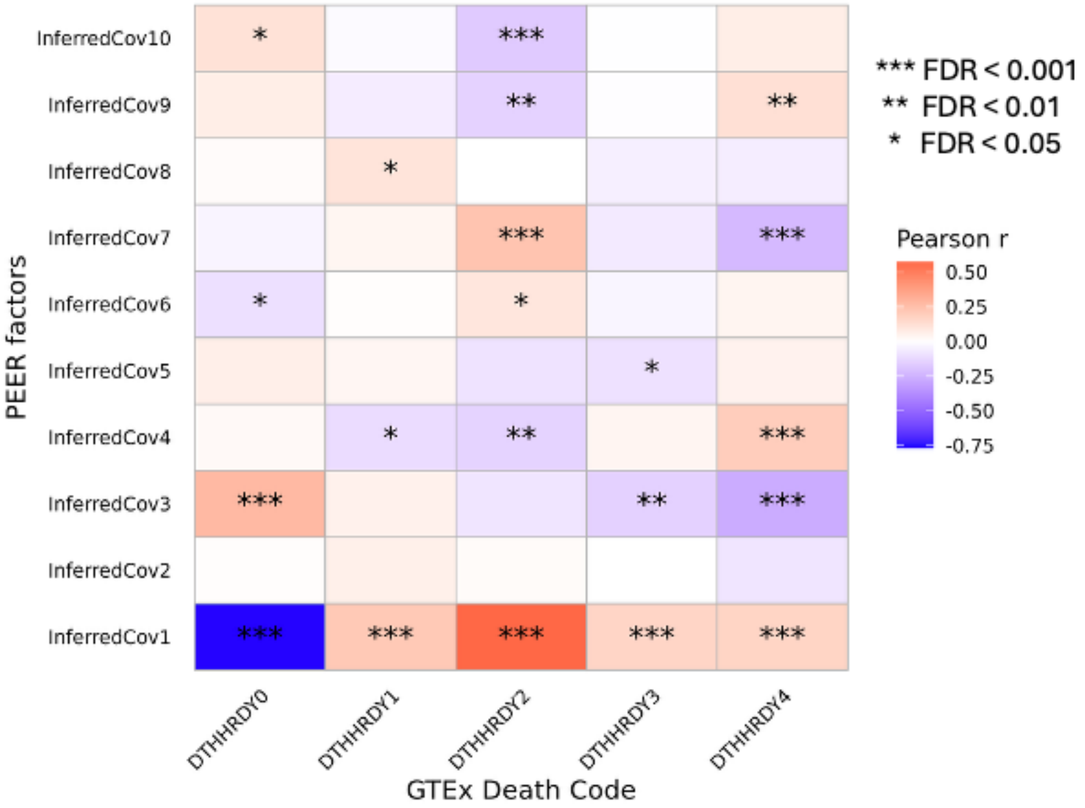


**Supplementary Figure 6. PEER factor analysis.** The top 10 PEER factors of gene expression measurements across individuals was highly correlated with metadata for GTEx cohort individuals, including death code which describes the cause of death. Hardy scale criteria defined by GTEx[25] as numeric value following "DTHHRDY": 0) Ventilator Case. All cases on a ventilator immediately before death. 1) Violent and fast death, i.e., deaths due to accident, blunt force trauma or suicide, where terminal phase estimated to be < 10 minutes; 2) Fast death of natural causes, i.e., sudden unexpected deaths of people who had been reasonably healthy or after a terminal phase estimated at < 1 hour (with heart attack being the most common cause); 3) Intermediate death, i.e., death after a terminal phase of 1 to 24 hours and patients who were ill but death was unexpected; 4) Slow death, i.e., death after a long illness, with a terminal phase longer than 1 day (commonly cancer or chronic pulmonary disease) and deaths that are not unexpected.

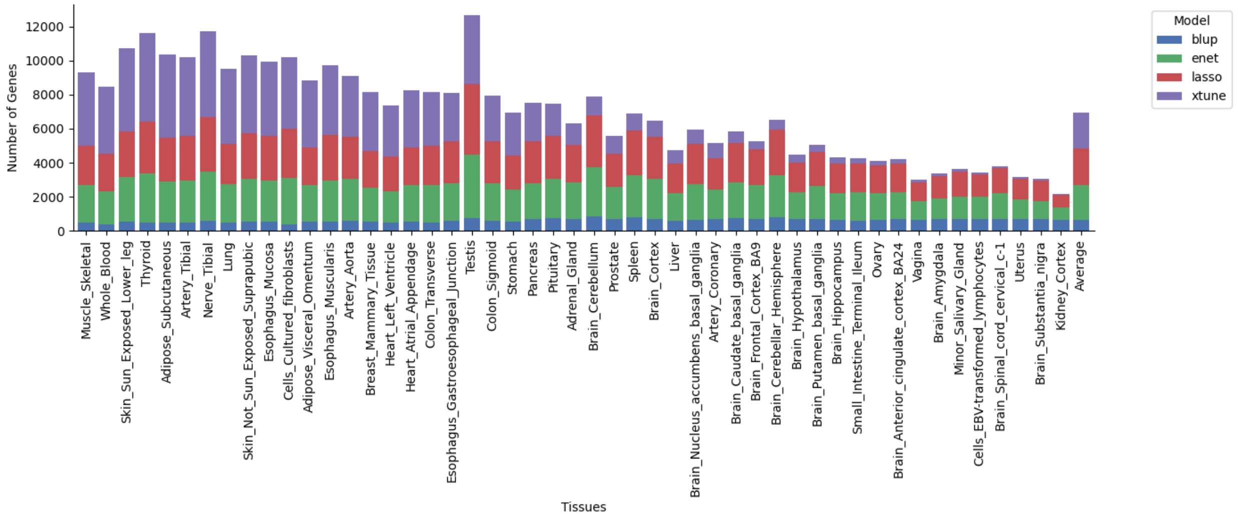


**Supplementary Figure 7. xtune LASSO comparison to other regularization schemes.** Across genes with predictive models ($R^2$ $p < 0.01$) across each of 49 tissues, we report how often each regularization approach (BLUP, elastic net (enet), LASSO, or xtune LASSO) resulted in the greatest CV $R^2$. Tissues are ordered in decreasing sample size from the left. In general, xtune LASSO performs the best for tissues with larger sample sizes, while LASSO and elastic net perform better for tissues with smaller sizes.

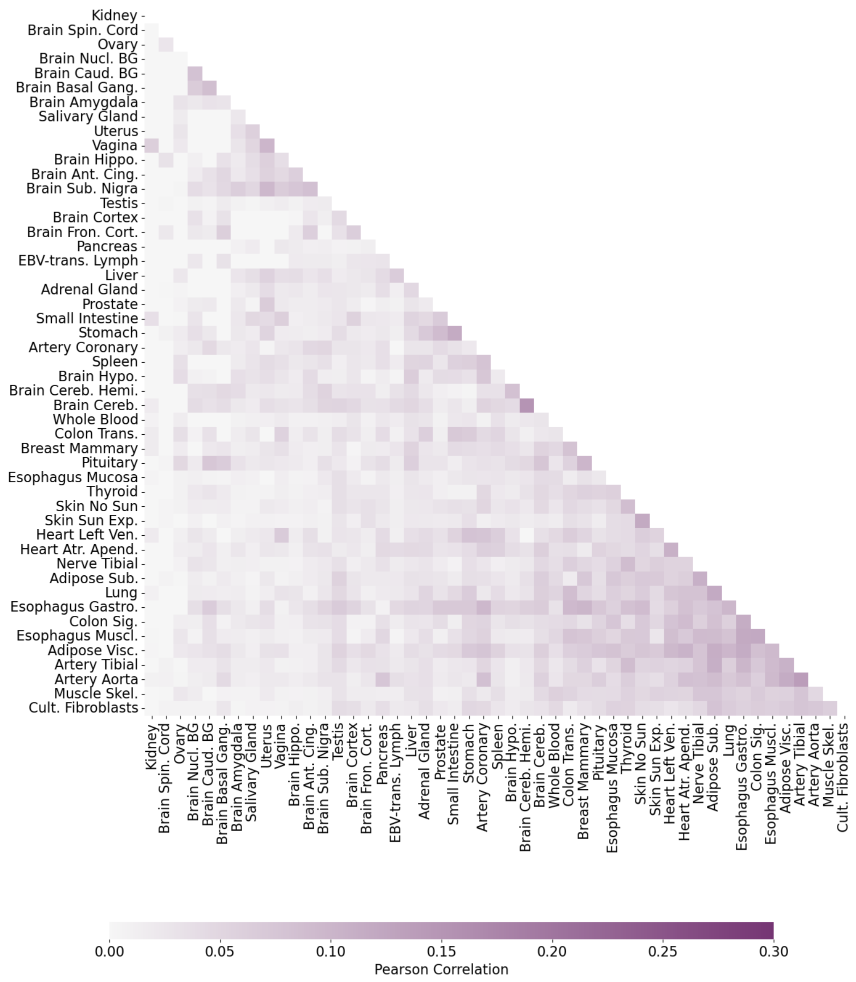


**Supplementary Figure 8. Cross-tissue correlations of *trans*-predicted gene expression.** Tissues are ordered according to hierarchical clustering of imputed expression using weighted trans-variants from EGRET models. Brain and reproductive tissues form a modest cluster in the top left, and chest tissues (esophagus and artery) form a modest cluster in the bottom right.

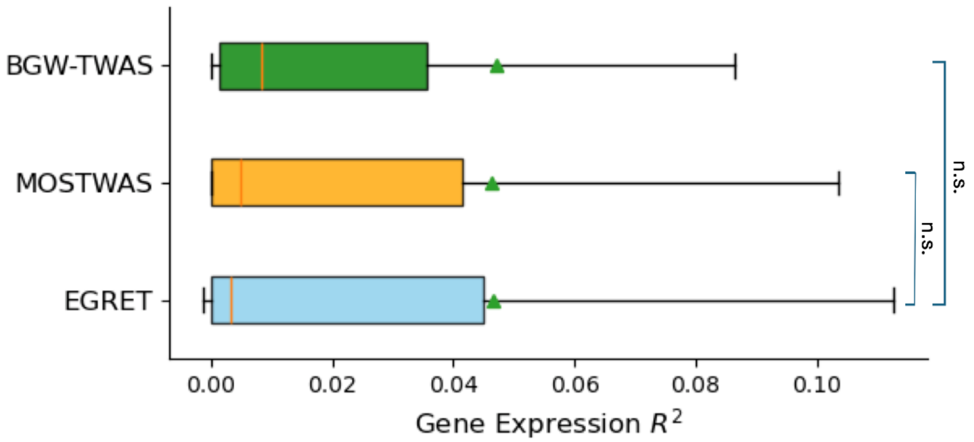


**Supplementary Figure 9. Distribution of gene model performances.** Comparison of $R^2$ distribution across non-cross-mappable genes using EGRET, BGW-TWAS, and MOSTWAS within GTEx whole blood. Green triangles denote means; red lines denote medians. n.s. indicates that there is no statistically significant difference between the means of these distributions.

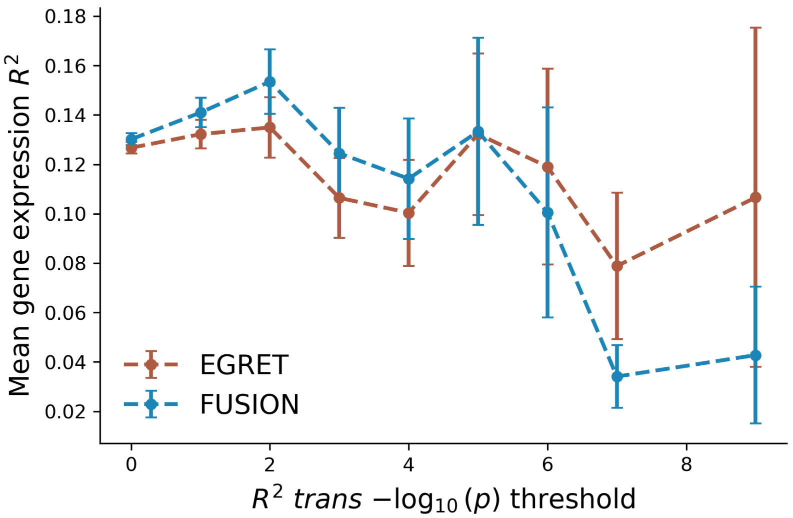


**Supplementary Figure 10. Comparison of FUSION and EGRET model $R^2$ in DGN cohort across different subsets of *trans*-heritable genes.** On the x-axis, we vary the p-value threshold to determine if a gene is *trans*-heritable according to the EGRET model (default $p < 0.01$, representing second dots/bars from the left). Bars indicate +/- 1 standard deviation from the average $R^2$ value across genes. From left to right, the size of the gene subset decreases, as there will be fewer genes with more highly significant $R^2_{trans}$ p-values.

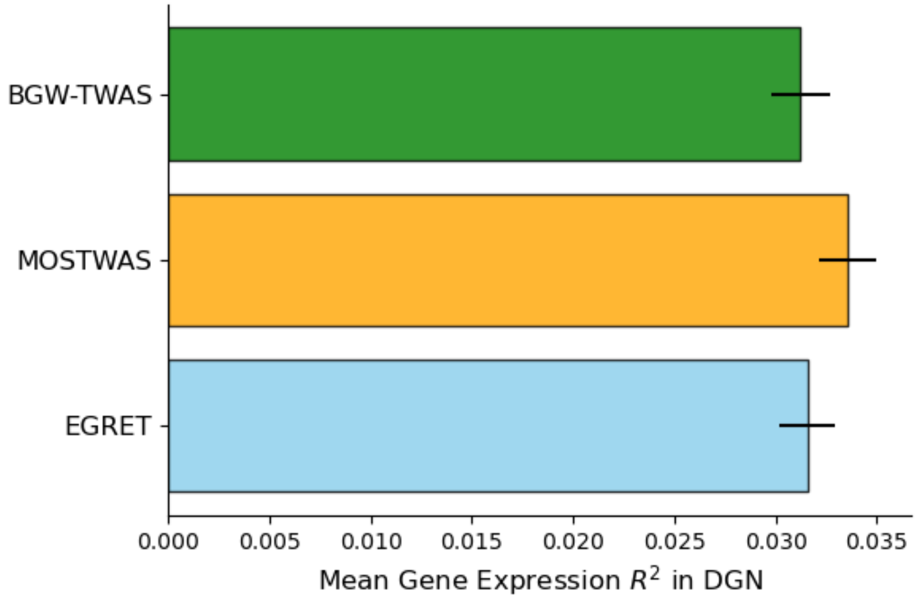


**Supplementary Figure 11. Distribution of gene model performances out of cohort.** Comparison of mean $R^2$ across non-cross-mappable genes using EGRET, BGW-TWAS, and MOSTWAS models trained on GTEx and applied to DGN whole blood tissue. Error bars represent one standard error.

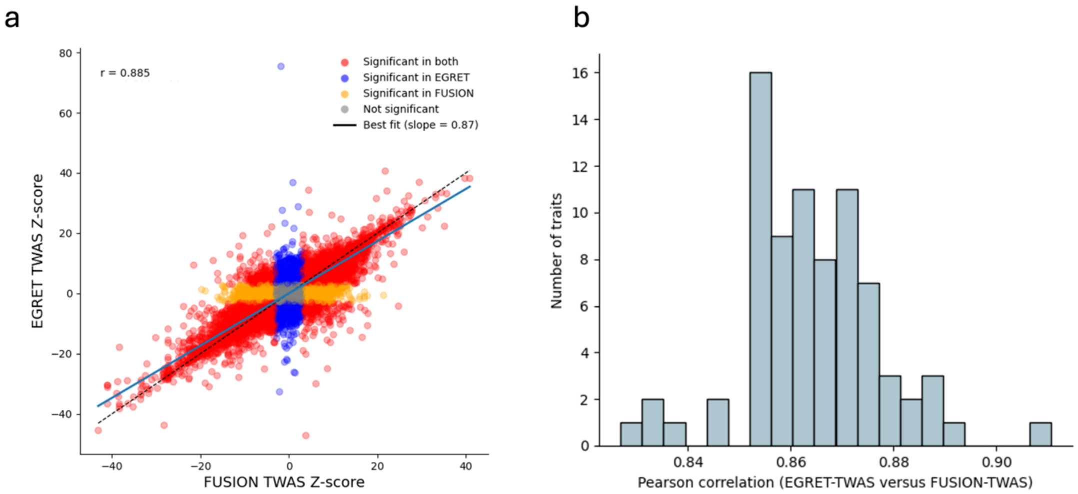


**Supplementary Figure 12. Concordance of EGRET-TWAS results with FUSION.** (a) Pearson correlation between EGRET-TWAS *z*-scores and FUSION-TWAS *z*-scores for a representative trait (platelet count) across all tissues in GTEx. Red points represent genes which are significant using both methods. Blue points are significant only in EGRET-TWAS and yellow are significant only in FUSION-TWAS. Gray dots are not significant in either method. (b) Histogram of Pearson correlations between EGRET-TWAS *z*-scores and FUSION-TWAS *z*-scores across all 78 diseases/traits analyzed. EGRET-TWAS and FUSION generally show strong concordance in TWAS *z*-score.

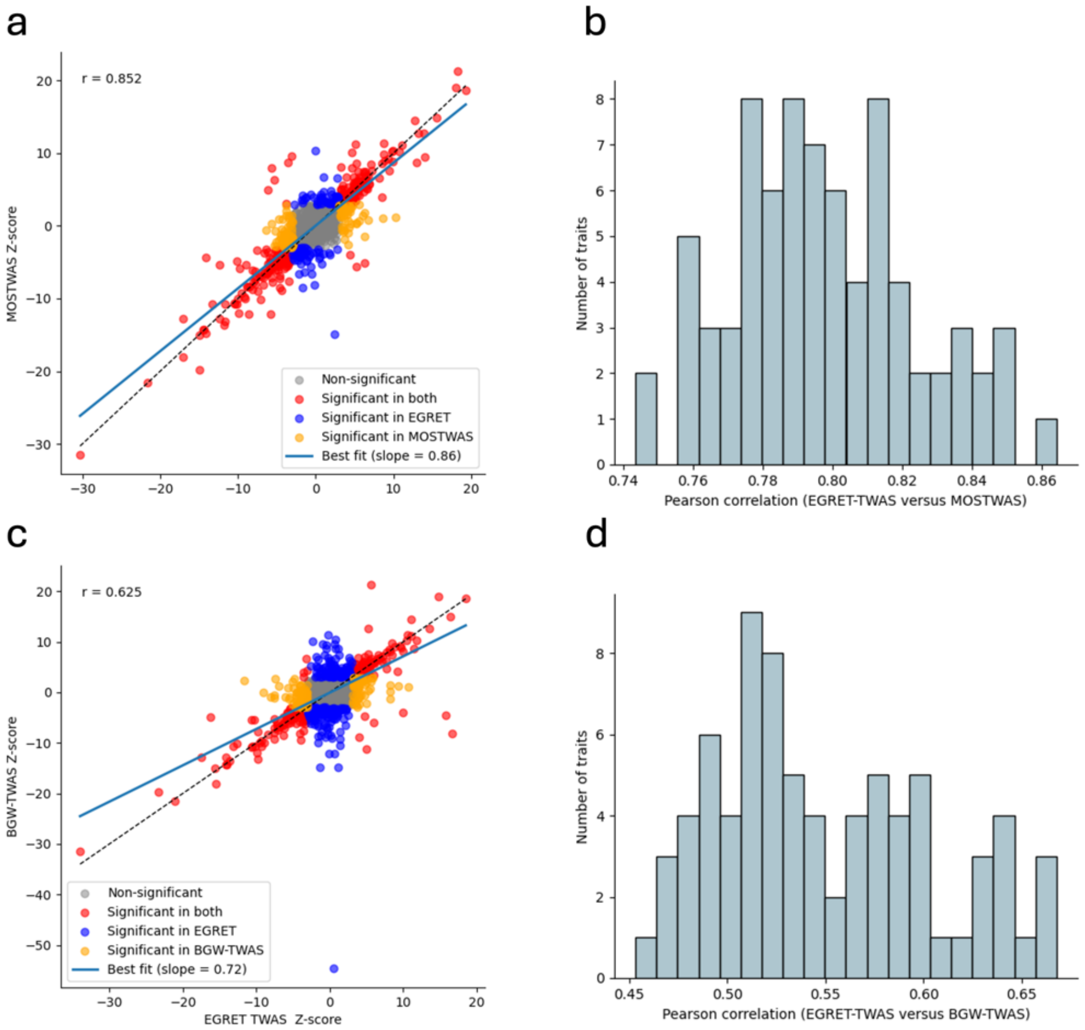


**Supplementary Figure 13. Concordance of EGRET-TWAS results with MOSTWAS and BGW-TWAS.** (a) Pearson correlation between EGRET-TWAS *z*-scores and MOSTWAS z-scores for a representative trait (platelet count) across non-cross-mappable genes in GTEx whole blood. (b) Histogram of Pearson correlations between EGRET-TWAS *z*-scores and MOSTWAS *z*-scores across the 78 diseases/traits analyzed. (c) Pearson Correlation between EGRET-TWAS *z*-scores and BGW-TWAS *z*-scores for platelet count across non-cross-mappable genes in GTEx whole blood. (c) Histogram of Pearson correlations between EGRET-TWAS *z*-scores and BGW-TWAS *z*-scores across the diseases/traits analyzed. Red points represent genes which are significant using both methods. Blue points are significant only in EGRET-TWAS and yellow are significant only in MOSTWAS (top) or BGW-TWAS (bottom). Gray dots are not significant in either method.

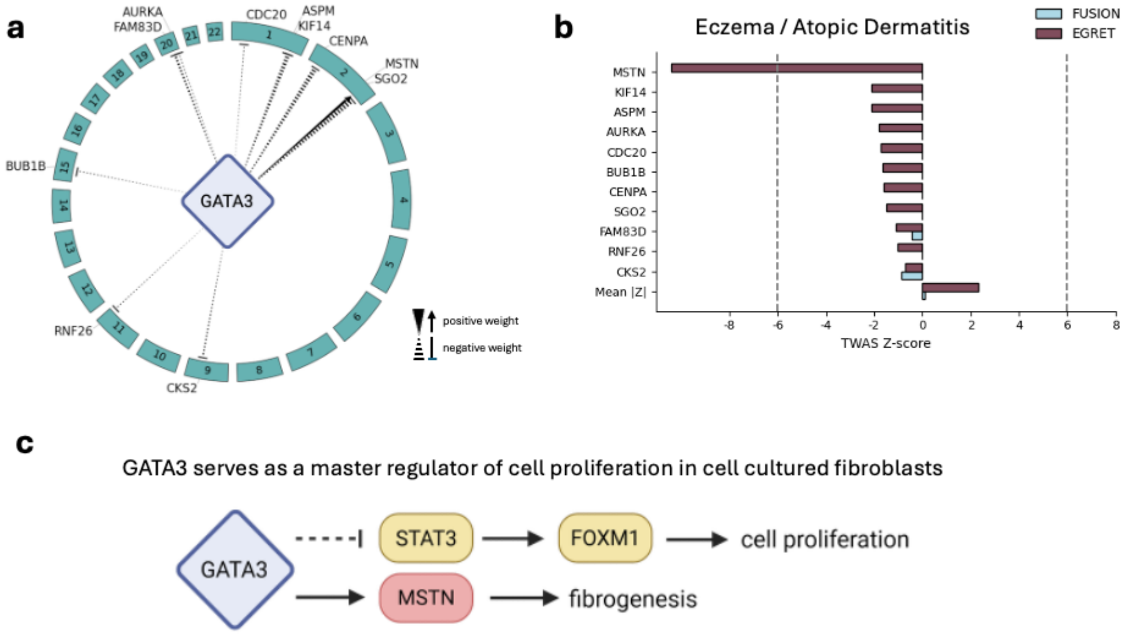


**Supplementary Figure 14. EGRET associates IRF4 regulatory network with eczema/atopic dermatitis.** (a) *IRF4* influences downstream genes across the genome. (b) FUSION and EGRET TWAS $z$-scores for eczema with genes in the *IRF4* module. (c) Schematic of established *IRF4* regulation potentially influencing genes involved in eczema/atopic dermatitis.

***Supplementary Note References***

122. Kim, K. W. *et al.* Genome-wide association study of recalcitrant atopic dermatitis in Korean children. *J. Allergy Clin. Immunol.* 136, 678-684.e4 (2015).

123. Shi, Q. *et al.* GATA3 suppresses human fibroblasts-induced metastasis of clear cell renal cell carcinoma via an anti-IL6/STAT3 mechanism. *Cancer Gene Ther.* 27, 726–738 (2020).

124. Mencalha, A. L., Binato, R., Ferreira, G. M., Du Rocher, B. & Abdelhay, E. Forkhead Box M1 (FoxM1) Gene Is a New STAT3 Transcriptional Factor Target and Is Essential for Proliferation, Survival and DNA Repair of K562 Cell Line. *PLoS ONE* 7, e48160 (2012).
